# Integrating Nowcasts into an Ensemble of Data-Driven Forecasting Models for SARI Hospitalizations in Germany

**DOI:** 10.1101/2025.02.21.25322655

**Authors:** Daniel Wolffram, Johannes Bracher, The RESPINOW Study Group, Melanie Schienle

## Abstract

Predictive epidemic modeling can enhance situational awareness during emerging and seasonal outbreaks and has received increasing interest in recent years. A common distinction is between nowcasting, which corrects recent incidence data for reporting delays, and forecasting, which predicts future trends. This paper presents an integrated system for nowcasting and multi-model short-term forecasting of hospitalizations from severe acute respiratory infections (SARI) in Germany (November 2023–September 2024). We propose a modular approach combining a statistical nowcasting model with various data-driven forecasting methods, including a time series model, a gradient boosting approach, and a neural network. These are combined into an ensemble approach, which achieves the best average performance. The resulting forecasts are overall well-calibrated up to four weeks ahead, but struggled to capture the unusual double peak which occurred during the test season. While the presented analysis is retrospective, it serves as a blueprint for a collaborative real-time forecasting platform for respiratory diseases in Germany (the *RESPINOW Hub*). We conclude with an outlook on this system, which was launched in the fall of 2024 and covers a broader range of data sources and modeling approaches.

## 1 Introduction

Predictive modeling of infectious diseases has received considerable attention in recent years. This was fueled by the public health crises of COVID-19 (e.g., Cramer et al. 2022; Bracher et al. 2021) and mpox (e.g., Bleichrodt et al. 2024), but general principles had been developed previously, most prominently for seasonal influenza (Reich et al., 2019). Disease forecasting is a broad field and three main types of predictive modeling tasks can be distinguished (Reich et al. 2022, see Figure 1).

- **Nowcasting** refers to the statistical correction of recent data points which are yet incomplete and subject to delayed additions. Nowcasts refer to recent rather than upcoming infection dynamics but are predictive in that they anticipate data revisions and reveal current trends.
- **Short-term forecasting** refers to unconditional predictions about the future course of an epidemic. Such predictions are feasible only for fairly short time periods, with appropriate prediction horizons depending on the type of indicator to predict.
- **Scenario predictions** are used to make statements about possible longer-term developments but are conditional on explicit assumptions that may or may not correspond to the future conditions encountered in the real world. For instance, scenarios may elucidate possible epidemic trajectories under various intervention strategies.

**Figure 1:**
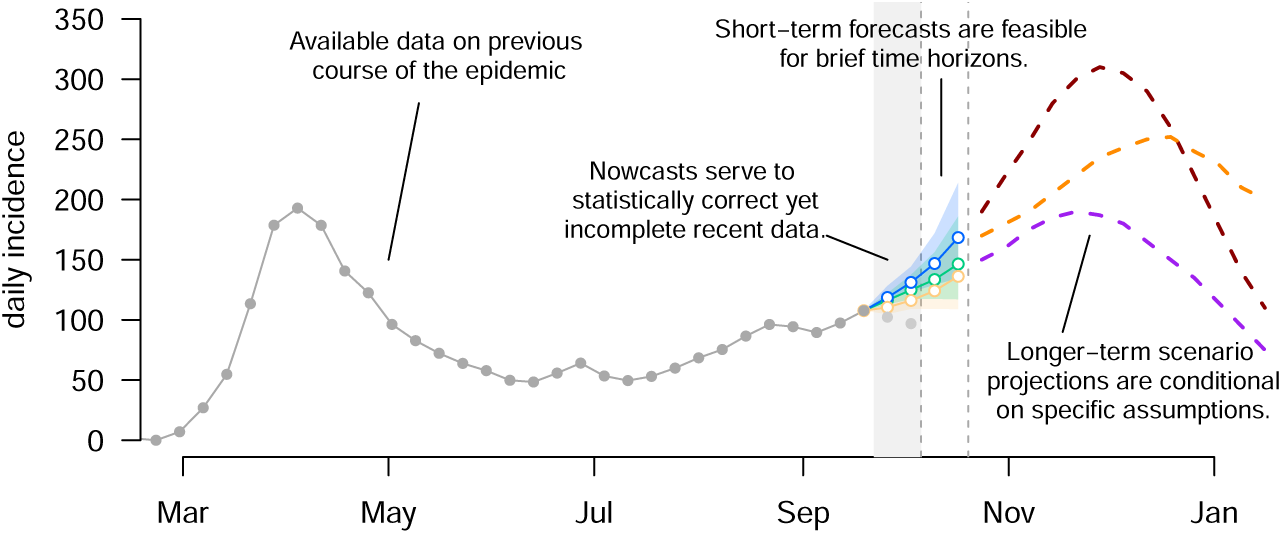
Distinguishing nowcasting, short-term forecasting, and scenario modeling of infectious diseases.

Scenario modeling is conceptually different from the other two settings in that it yields statements about hypothetical settings rather than verifiable predictions. Nowcasting and short-term forecasting, on the other hand, ultimately boil down to the same task, namely probabilistic statements about disease incidence at various points in time. The availability of preliminary incidence values makes this more straightforward in nowcasting, but there is a natural transition between the two tasks. Somewhat surprisingly, nowcasting and short-term forecasting are rarely addressed jointly (an exception being Beesley et al. 2022).

For all three tasks, there is a growing consensus that multi-model approaches are particularly suitable. The presence of multiple distinct models enables more realistic assessments of the predictive uncertainty and can be the basis for ensemble forecasts, which have often been found to be more robust. Multi-model forecasting is often conducted in collaborative *Hubs* which collect contributions by participating teams. Recent efforts include e.g., the US COVID-19 Forecast and Scenario Modeling Hubs (Cramer et al., 2022; Howerton et al., 2023), the European COVID-19 Forecast Hub (Sherratt et al., 2023) and the German COVID-19 Hospitalization Nowcast Hub (Wolffram et al., 2023). While the latter was explicitly limited to nowcasting, the forecasting platforms circumvented the nowcasting problem in various ways. As detailed e.g., in Bracher et al. (2021), this can be done by aggregating incidence counts according to the date of report rather than e.g., symptom onset. Alternatively, the most recent data points can be removed entirely to avoid dealing with delays, as done e.g., in a French hospitalization forecasting system (Paireau et al., 2022). Both approaches, however, blur or even discard valuable information on recent developments.

In this paper, we present a combined nowcasting and multi-model short-term forecasting system for hospitalizations due to severe acute respiratory infections (SARI) in Germany (November 2023–September 2024). Unlike forecast targets based on reporting dates, SARI hospitalizations are recorded by the date of hospital admission, making them susceptible to reporting delays and revisions, which necessitates special handling of the most recent, yet incomplete, data points. To this end, we develop a modular approach, where, rather than integrating a nowcasting step into each individual forecasting model, it is split off and handled by a separate statistical model. The resulting probabilistic nowcasts are fed back into a variety of forecasting methods. This includes a statistical time series model, a gradient boosting approach, and a neural network, which are moreover combined into an ensemble. The modular approach is practical as it avoids integrating nowcasting steps separately into conceptually diverse forecasting techniques. Moreover, it is helpful when historical data snapshots are not available for the entire period of interest; indeed, in our application, the first available data snapshot is from a release in September 2023, but the contained time series data reaches back to 2014. Splitting the training of nowcasting and forecasting models facilitates exploiting these disparate data sets in a straightforward manner. While the current paper is retrospective in nature, the developed approaches serve as a blueprint for a collaborative real-time nowcasting and forecasting system of infectious disease spread in Germany. As we will detail in Section 5, the so-called *RESPINOW Hub* was launched in fall 2024, and a prospective evaluation study on the 2024/25 season has recently been preregistered (Bracher and Wolffram, 2024).

A specific challenge in our application arises from the fact that the COVID-19 pandemic not only added to the general respiratory disease burden but also strongly impacted the dynamics of other respiratory diseases (see, e.g., Buchholz et al. 2023). This is true for the years 2020–2022 when the associated non-pharmaceutical interventions largely stopped the spread of other respiratory diseases, but also the following period, when the immunity landscape was considerably different from earlier years. We will compare different approaches to using historical data from these periods for model fitting.

We find our forecasting models to be generally well-calibrated, though with rather wide uncertainty intervals surrounding peak weeks, and some difficulty in dealing with the double peak occurring in the test season. Similarly to previous studies, an ensemble approach achieves the best overall performance. Including a nowcasting step clearly improves forecasts relative to a procedure where the most recent data point is simply discarded. Indeed, the loss in forecasting performance relative to a hypothetical setting where the data are not subject to reporting delays turns out to be minor. This leads us to recommend the inclusion of nowcasting steps in infectious disease forecasting systems.

The remainder of the paper is structured as follows. Section 2 provides background information on SARI hospitalizations in Germany. In Section 3, we define the nowcasting and forecasting targets and present the methods employed for both tasks. Particular attention will be paid to the question of how to feed nowcast information into forecasting models while accounting for the arising uncertainties. In Section 4, we evaluate the resulting probabilistic forecasts visually and with a variety of metrics. In Section 5 we provide an outlook on the *RESPINOW Hub* before concluding with a discussion in Section 6.

## 2 The SARI hospitalization incidence

### 2.1 Definition and description

Respiratory disease activity in Germany is subject to a multitude of surveillance systems (see also Section 6). In the present paper, we are concerned with the hospitalization incidence for *severe acute respiratory infections* (SARI). Since fall 2014, data on such hospitalizations have been collected in the *ICOSARI* system operated by Robert Koch Institute (RKI; Buda et al. 2017; Tolksdorf et al. 2022). They are publicly accessible via the RKI GitHub repository (https://github.com/robert-koch-institut/SARI-Hospitalisierungsinzidenz). The SARI hospitalization incidence is a *syndromic indicator*, i.e., the case definition is based on the symptoms patients present rather than laboratory testing for a specific pathogen. Specifically, a set of ICD-10 diagnostic codes (J09–J22) is used, see Buda et al. (2017) for details. Data collection is carried out via a sentinel system that includes roughly 70 hospitals in 13 of the 16 German federal states. The system covers around 6% of all hospitalizations occurring in Germany. Based on an estimation of the catchment population covered by the sentinel sites, the SARI hospitalization incidence per 100,000 inhabitants can be estimated. Estimates in a weekly resolution (with weeks starting on Mondays) are made available both unstratified (00+) and for six age groups (0–4, 5–14, 15–34, 35–59, 60–79, 80+). In this paper, we rescale the estimated incidence to absolute count values.

The pooled and age group-wise incidence time series for the period 2014–2024 are displayed in Figure 2. Seasons we consider substantially affected by the acute phase of the COVID-19 pandemic are delimited by dashed vertical lines. Colors moreover indicate the split into training, validation, and test periods, see Section 4.2.4 for details. Especially in the age groups 05–14, 15–34, and 35–59, the test season displays rather unusual patterns, with consistently high incidences even in late spring and summer. In the very young and old age groups, this is less pronounced. As these age groups feature higher absolute numbers, the pooled incidence shown in the left panel likewise shows a more typical seasonal course.

**Figure 2:**
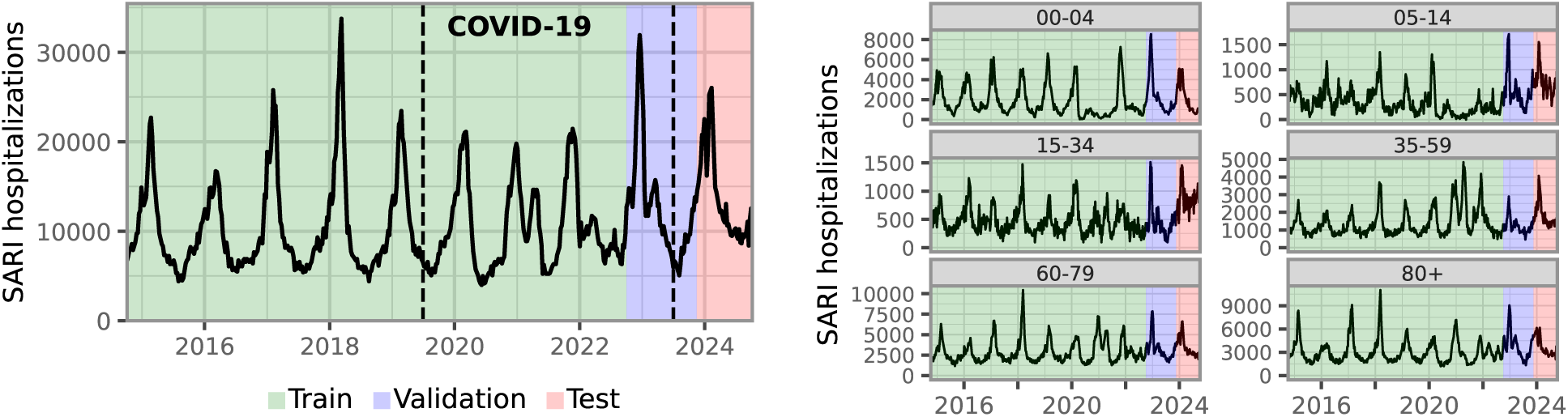
Time series of weekly SARI hospitalizations in Germany, 2014–2024. Colors indicate the split of the data into training, validation, and test data; see details in Section 3.3.2. The portion labeled “COVID-19” is only included in the training set for part of our model specifications.

We note that two of the considered forecasting models (LightGBM and TSMixer; Section 3.3) use an auxiliary data set on weekly outpatient consultations for acute respiratory infections (ARI). Details on this data set are provided in Supplement B and a visualization is shown in Figure S1.

### 2.2 Data revisions and reporting delays

Like many epidemiological indicators, the SARI hospitalization incidence is subject to retrospective data revisions. Typically, the numbers are corrected upwards as additional hospitalizations are reported with a delay. To assess the impact of reporting delays, archives of historical data snapshots are necessary. The public RKI GitHub repository contains such snapshots back to the data release on 28 September 2023. Prior to this date, PDF reports were made available which enabled the reconstruction of snapshots at the aggregate level back to early 2023 (though not for the different age groups).

As illustrated in the left panel of Figure 3, reporting delays lead to an artificial dip towards the end of the incidence time series data available in real time. Once data points have been completed over the following weeks, this dip disappears, and the actual trend becomes visible. For the SARI data, corrections become largely negligible after three weeks. The right panel shows the completeness of the data zero to four weeks after the initial release, by data release week. It can be seen that initial data releases on average contain roughly 75% of the hospitalizations (or, put differently, initial values are corrected upwards by roughly a third). Initial reporting completeness fluctuates somewhat over time. Between Christmas and New Year, no release occurs, meaning all hospitalizations from this period are reported with a delay.

**Figure 3:**
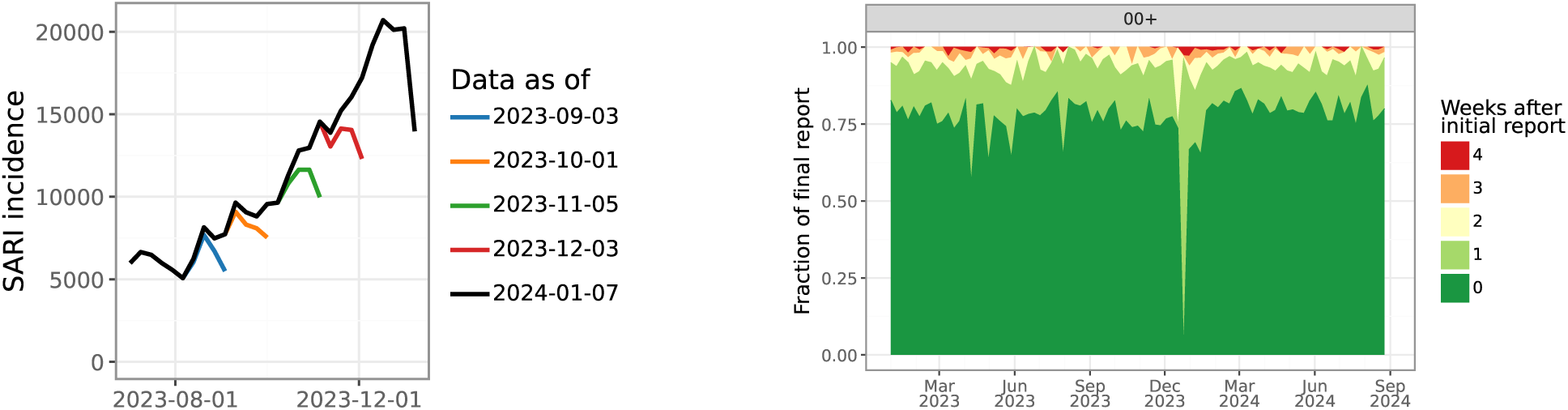
Left: Illustration of data revisions in the SARI hospitalization incidence. Time series as available on different dates are shown in different colors, overlaid with more complete data in black. The apparent downward trend in the initial data versions is replaced by a continued upward trend in the revised data. Right: Completeness of SARI hospitalization data zero to four weeks after the first release, per week (2023–2024). In alignment with the nowcasting target definitions (see Section 3), we only consider delays up to four weeks.

## 3 Methods

### 3.1 Definition of nowcasting / forecasting tasks

Nowcasting addresses the statistical correction of partial/preliminary data. Real-time surveillance data are commonly completed or revised retrospectively. This is because by the time a new version of a surveillance data set is published, not all relevant reports will already have been received by the organization curating the data. Later versions of the data set will be updated with additional information. Nowcasting typically, but not necessarily, corresponds to upward correction of data to account for delayed reports. Forecasting concerns the future epidemiological development and thus time points for which not even partial data is currently available.

We generate nowcasts and forecasts in a weekly rhythm for the time period from 16 November 2023 through 12 September 2024, following the data release schedule on Thursdays. We skipped Thursday 28 December 2024 as no data release was available. This *test period* is highlighted in red in Figure 2. Counting from the day of data release (Thursday), the week ending on the preceding Sunday is indexed as *horizon* or *lead time* 0 weeks. Nowcasts, i.e., corrections of available preliminary data for e.g. reporting delays, are produced for weeks −3 through 0. Forecasts are generated for horizons 1 through 4. All predictions are generated both for the total weekly number of SARI hospitalizations on the national level (aggregated across all ages) and stratified per age group. We note that the available SARI hospitalization incidence is actually only an estimate (see previous section). In practice, we neglect any uncertainty attached to these estimates and simply treat the estimates as the observable prediction target.

In the presence of data revisions, the definition of the prediction targets requires specific care. Based on experience from previous work (Wolffram et al., 2023), we define the final data version against which both nowcasts and forecasts are evaluated via a maximum reporting delay of *D* = 4 weeks. For each week, the respective data point used in the evaluation is thus set to the value available after four weeks of revisions (i.e., as published four weeks after the first data release containing a value for the respective week). This definition has the advantage of providing a well-defined target, with all observations in the evaluation period given the same amount of time for revisions. It is, however, unusual in that the time series used for evaluation is not identical to any specific public data release.

For each nowcast or forecast horizon, we collect predictive quantiles at levels 2.5%, 10%, 25%, 50%, 75%, 90%, and 97.5%. This storage format corresponds to that of various Forecast Hubs established during the COVID-19 pandemic (Cramer et al., 2022; Wolffram et al., 2023). While it brings some constraints with it, we follow this convention as the present analysis aims to support the development of a new collaborative forecasting platform in Germany (see Section 5).

### 3.2 Nowcasting method and the coupling of nowcasting and forecasting

We separate the nowcasting and forecasting step and use a separate nowcasting model, which provides input for several forecasting models. While it may seem desirable to integrate nowcasting directly into each forecasting method, in practice this is hard to accommodate in many cases. Another challenge is the fact that data snapshots could only be partly recovered. We therefore split off the nowcasting from the forecasting task.

For nowcasting, we suggest a method that is based on the simpleNowcast first discussed in Wolffram et al. (2023, Supplementary Section E). It combines a straightforward multiplication factor scheme with a parametric approach to estimate predictive uncertainty from past nowcast errors. Despite its simplicity, the approach showed performance comparable to more sophisticated approaches in Wolffram et al. (2023). In the present application, we need to adapt the original approach from daily to weekly data releases, which actually simplifies the technique as data release and now/forecast schedules share the same frequency. The chosen simple format of the suggested nowcast technique has the key advantage that it allows to easily deal with very limited or missing information on (age) strata that characterize our data.

#### 3.2.1 Point nowcast

Denote by *X_t,d_, d* = 0, …, *D* the number of hospitalizations for week *t* which appear in the data set with a delay of *d* ≥ 0 weeks. In our applied setting, delay *d* = 0 means that a hospitalization from the week ending on a given Sunday was already included in the data release from the following Thursday. Note that we only consider hospitalizations reported with a maximum of *D* weeks (in our application *D* = 4). We now denote by

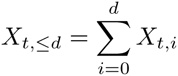

the number of hospitalizations reported for week *t* with a delay of at most *d* weeks, implying that *X_t_* = *X_t,_*_≤*D*_. Conversely, for *d < D*

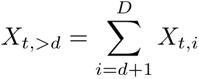

is the number of hospitalizations still missing after *d* weeks.

In the following, we write *X_t_* etc. for a random variable, *x_t_* for the corresponding observation and *x̂_t_* for an estimated/imputed value. The hospitalizations per week and reporting delay as available at a given data release time *t*\* can be arranged into a *reporting triangle*, see Table 1.

**Table 1:** Illustration of the reporting triangle for time *t*\* and *D* = 4. Quantities known at time *t*\* are shown in black, yet unknown quantities are shown in

| day | $d = 0$ | $d = 1$ | $d = 2$ | $d = 3$ | $d = 4$ | total |
| --- | --- | --- | --- | --- | --- | --- |
| 1 | $x_{1,0}$ | $x_{1,1}$ | $x_{1,2}$ | $x_{1,3}$ | $x_{1,4}$ | $x_1$ |
| 2 | $x_{2,0}$ | $x_{2,1}$ | $x_{2,2}$ | $x_{2,3}$ | $x_{2,4}$ | $x_2$ |
| ⋮ | | | | | | |
| $t^* - 5$ | $x_{t^*-5,0}$ | $x_{t^*-5,1}$ | $x_{t^*-5,2}$ | $x_{t^*-5,3}$ | $x_{t^*-5,4}$ | $x_{t^*-5}$ |
| $t^* - 4$ | $x_{t^*-4,0}$ | $x_{t^*-4,1}$ | $x_{t^*-4,2}$ | $x_{t^*-4,3}$ | $x_{t^*-4,4}$ | $x_{t^*-4}$ |
| $t^* - 3$ | $x_{t^*-3,0}$ | $x_{t^*-3,1}$ | $x_{t^*-3,2}$ | $x_{t^*-3,3}$ | $x_{t^*-3,4}$ | $x_{t^*-3}$ |
| $t^* - 2$ | $x_{t^*-2,0}$ | $x_{t^*-2,1}$ | $x_{t^*-2,2}$ | $x_{t^*-2,3}$ | $x_{t^*-2,4}$ | $x_{t^*-2}$ |
| $t^* - 1$ | $x_{t^*-1,0}$ | $x_{t^*-1,1}$ | $x_{t^*-1,2}$ | $x_{t^*-1,3}$ | $x_{t^*-1,4}$ | $x_{t^*-1}$ |
| $t^*$ | $x_{t^*,0}$ | $x_{t^*,1}$ | $x_{t^*,2}$ | $x_{t^*,3}$ | $x_{t^*,4}$ | $x_{t^*}$ |

We consider data as available in week *t*\* and aim to obtain point nowcasts 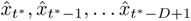, i.e., for all observations which in week *t*\* are still incomplete. We start by setting

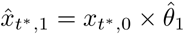

with a multiplication factor

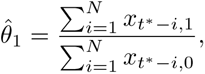

obtained from *N* preceding rows of the triangle. Here, the estimation window size *N < t*\* is chosen by the user and serves to restrict the estimation to fairly recent data. In practice we use *N* = 15, implying that snapshots from at least the last 15 weeks are needed. Following the same principle, we compute

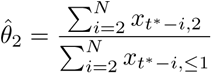

and use it to impute

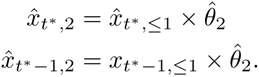

Here, we use the 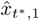 imputed in the first step to compute

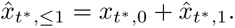

The same procedure is applied to all other missing values of the reporting triangle, which we fill from the left to the right and the bottom to the top.

For *d* = 0, …, *D* − 1 we then sum over relevant entries of the imputed reporting triangle to obtain point nowcasts

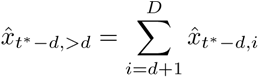

for the hospitalizations from week *t*\* − *d* that are still to be reported. Point nowcasts for the total numbers result as

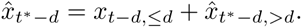

A slightly more formal explanation on how this relates to the estimation of a delay distribution from censored observations can be found in Wolffram et al. (2023). We note that this scheme would require some adaptations to deal with zeros in the reporting triangle, but these do not occur in our setting.

#### 3.2.2 Nowcast uncertainty

We now describe how to extend these point nowcasts to probabilistic nowcasts based on past nowcast errors. To this end we need to slightly extend notation and write

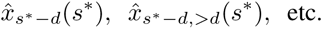

for nowcasts referring to week *s*\* − *d* and generated based on data as available in week *s*\*. As the uncertainty in the nowcasts only stems from the hospitalizations still to be added to the record we focus on the 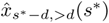 in the following.

Again consider the generation of nowcasts in week *t*\*. To quantify the prediction uncertainty we start by computing 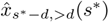 for *s*\* = *t*\* − *D, …, t*\* − *M* and *d* = 0, …, *D* − 1. In practice, we use *M* = 15. Note that to perform all these computations, data snapshots from at least *N* + *M* (hence in our case 30) past weeks are needed.

For each horizon *d* = 0, …, *D* − 1 we then assume that

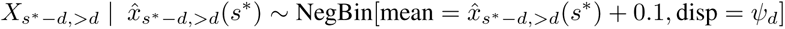

independently for each *s*\* = *t*\* − *D, …, t*\* − *M*. An estimate 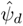 for the dispersion parameter is obtained via maximum likelihood inference. The addition of a small value of 0.1 serves to ensure well-definedness of the negative binomial distribution if 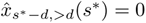. In practice, we add a little tweak to also be able to include partial observations from *s*\* = *t*\* − 1, …, *t*\* − *D* + 1, see Wolffram et al. (2023) for details. Our nowcast distribution for 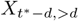 is then simply

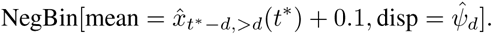

The corresponding distribution for the total count *X_t_*∗ results from shifting this distribution by the known count 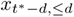.

We note that if *x_t,_*_0_ = 0 for a given week, i.e., there are no initial releases, we remove the respective row from the reporting triangle. This serves to catch weeks like the Christmas week when data releases are paused.

We chose this very simple methodology because it is straightforward to adapt to a particularity of the nowcasting task at hand. In practice we encounter the problem that historical data snapshots are only available for the total SARI hospitalization incidence, but not the age-stratified time series (see Section 2.2). To nonetheless produce nowcasts at the stratified level we simplifyingly assume that the reporting delay distribution is identical across strata. The parameter estimates 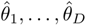 are thus estimated from the pooled reporting triangles. The estimated overdispersion parameters 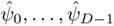 are likewise borrowed from the pooled fits.

#### 3.2.3 Coupling of nowcasting and forecasting

For coupling the nowcast with the forecasting models, we propose the following model-agnostic approach to propagate nowcast uncertainty into forecasts (illustrated in Figure 4):

1. Generate nowcast distributions for horizons −3 through 0 using a separate nowcasting model. For each horizon, quantiles at *N* = 39 different levels 0.025, 0.05, …, 0.95, 0.975 are generated.
2. Translate these into 39 sample paths by assembling the predictive quantiles at identical levels for the four horizons.
3. Feed each of these paths into the employed forecasting model to generate predictive distributions for horizons 1 through 4 (depending on the method these are samples or parametric distributions).
4. Combine these predictive distributions by aggregating samples or averaging probability mass functions with linear pooling.

**Figure 4:**
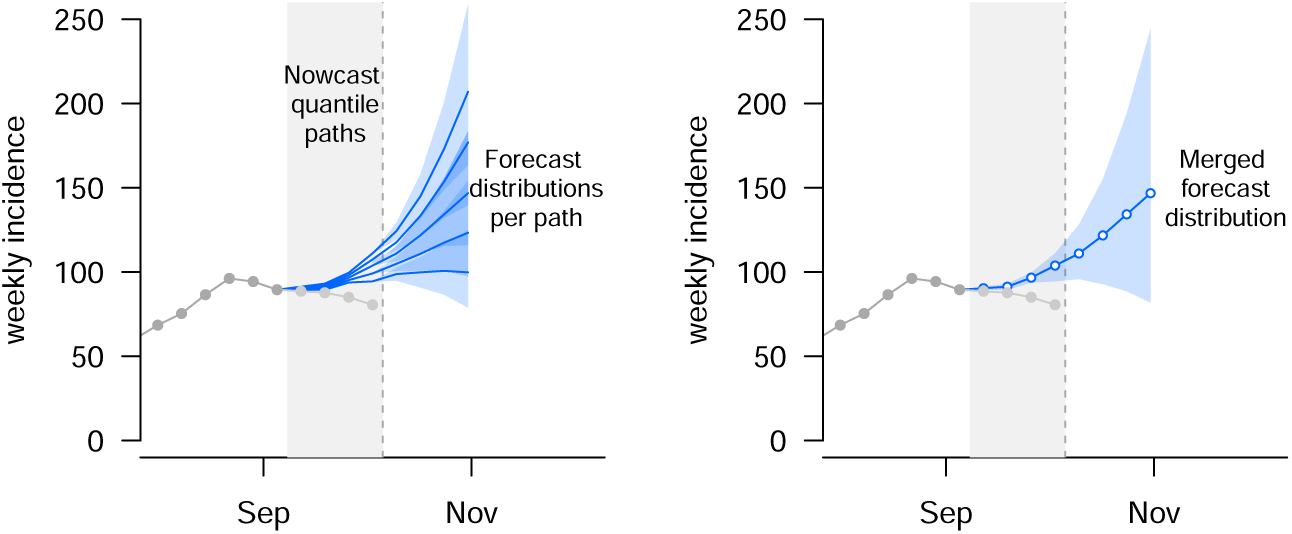
Illustration of coupling between nowcasting and forecasting. A set of nowcast sample paths (blue lines in grey shaded area) are generated. Each of these is def into a forecasting model to obtain predictive distributions for horizons 1 through 4. Results are then aggregated into overall forecast distributions via a linear pool (right panel).

Step 2 is arbitrary in a sense as the distributions our nowcasting model returns for the various horizons are purely univariate and nothing is known about the dependence structure. However, in practice, the corrections at horizons −3 through −1 are minor and unlikely to have a major impact on predictions. It is therefore not crucial how exactly the nowcast paths are formed.

### 3.3 Forecasting methods

As SARI is not caused by an individual pathogen, it is not straightforward to model its dynamics mechanistically using classic compartmental (SIR-type) models. However, the SARI indicator is characterized by strong autocorrelations and, at least up to the COVID-19 pandemic, rather stable seasonal patterns. We therefore develop a suite of statistical models to exploit these regularities. As detailed in the following sections, some models can moreover exploit multivariate patterns across age groups (such as respiratory diseases often spreading from the younger to the older age groups) and information contained in auxiliary data streams.

#### 3.3.1 Endemic-epidemic modeling: hhh4

The *endemic-epidemic* or hhh4 model (after the associated function in the R package surveillance, Meyer et al. 2017) is a statistical time series model tailored to infectious disease surveillance data. While in principle it is capable of reflecting dependence structures across space or age groups, in our setting a simple univariate formulation for each stratum proved most robust. Denoting the incidence value (as absolute count value) in week *t* by *X_t_*, the model is then defined as

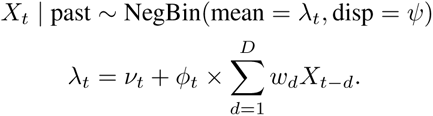

Here, the negative binomial distribution is parameterized by its mean *λ_t_* and an overdispersion parameter *ψ*. Following Bracher and Held (2022), we use geometrically decaying weights *w_d_*, while accounting for yearly seasonal variation via time-varying parameters. In the model for the pooled time series we used the standard formulation

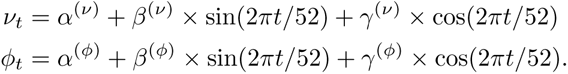

For the age-stratified forecasts, we further simplified this model and removed the intercept term *ν_t_*(i.e., set it to zero). Already during the training period, retrospective forecasts from models including the intercept did not adapt well adapt to changed magnitudes of incidence compared to earlier seasons. Especially in the age groups 05–14 and 35–59, this led to forecasts which were poorly aligned with the preceding data points. By removing the intercept this could be mitigated to a large degree.

Inference is conducted using maximum likelihood and predictions are obtained in a simple plug-in manner. Predictive first and second moments can be computed analytically for all forecast horizons (Bracher and Held, 2022), and matching negative binomial distributions are used to obtain quantiles.

The model fits are updated each week based on all historical data available (or, in a sensitivity analysis, excluding seasons strongly affected by the COVID-19 pandemic). Note that this also includes the corrected data points generated in the nowcasting step (see Section 3.2). Unlike the methods described in the two following subsections, no validation set is required, meaning that the distinction between the green and blue sections in Figure 2 is not relevant here. No additional data inputs other than the SARI incidences are used.

#### 3.3.2 Gradient boosting: LightGBM

LightGBM (Light Gradient Boosting Machine) is a gradient boosting framework designed for high-performance machine learning tasks (Ke et al., 2017). It builds decision tree ensembles sequentially, where each tree corrects the errors of the previous ones, enabling the model to capture complex patterns in the data. Its ability to efficiently handle large datasets, categorical variables, and missing values makes it versatile for a wide range of applications. In time series forecasting, LightGBM can effectively model relationships within multivariate data and incorporate exogenous variables. In the M5 forecasting competition (Makridakis et al. (2022)), the model was one of the top models for predicting retail sales across multiple products and stores.

For our analysis, the model was retrained each week based on the historical data available and implemented in a multivariate fashion, allowing simultaneous prediction of all targets (i.e., different age groups and the national level). Weekly ARI numbers (see Supplement B) were included as a covariate. In addition to the lagged values of these two time series, the calendar week and the month of the subsequent week were also incorporated as input features. The last few observations that would remain incomplete in a real-time setting were excluded from the training process. They were subsequently replaced by nowcast paths to compute the forecasts as described in Section 3.2. Concerning hyperparameter selection (executed with W&B by Biewald 2020), after an initial random search to identify promising regions, an exhaustive grid search was performed over the refined hyperparameter space described in Supplementary Table S1. To reduce computational requirements, the model was trained once on the training dataset and evaluated across all dates in the validation period (highlighted in green and blue in Figure 2). Due to the non-deterministic nature of the training process, we conducted training using 10 different random seeds and averaged the forecasts from these models (i.e., the predictive quantiles at each level) to obtain more robust results.

#### 3.3.3 Deep learning model: TSMixer

The TSMixer architecture (as introduced in Chen et al. (2023)) is a fully connected neural network specifically designed for time series forecasting. It utilizes a sequential mixing layer strategy that enables the model to capture both temporal dependencies and cross-feature interactions. As illustrated in Figure 5, the mixing layers are applied sequentially: first across the time dimension to model temporal patterns and then across the feature dimension to capture relationships between different variables. This approach allows the model to learn complex and non-linear relationships within the time series data. Compared to state-of-the-art transformer-based models, TSMixer often exhibits a simpler architecture, making it more computationally efficient and easier to train. Despite its relative simplicity, TSMixer has demonstrated competitive performance on a wide range of time series forecasting benchmarks, suggesting that its sequential mixing layer strategy is an effective approach for modeling temporal data. The model’s ability to handle multivariate time series, as well as its potential for incorporating exogenous variables, makes it a versatile tool for various time series forecasting applications, such as infectious disease forecasting in our setting.

**Figure 5:**
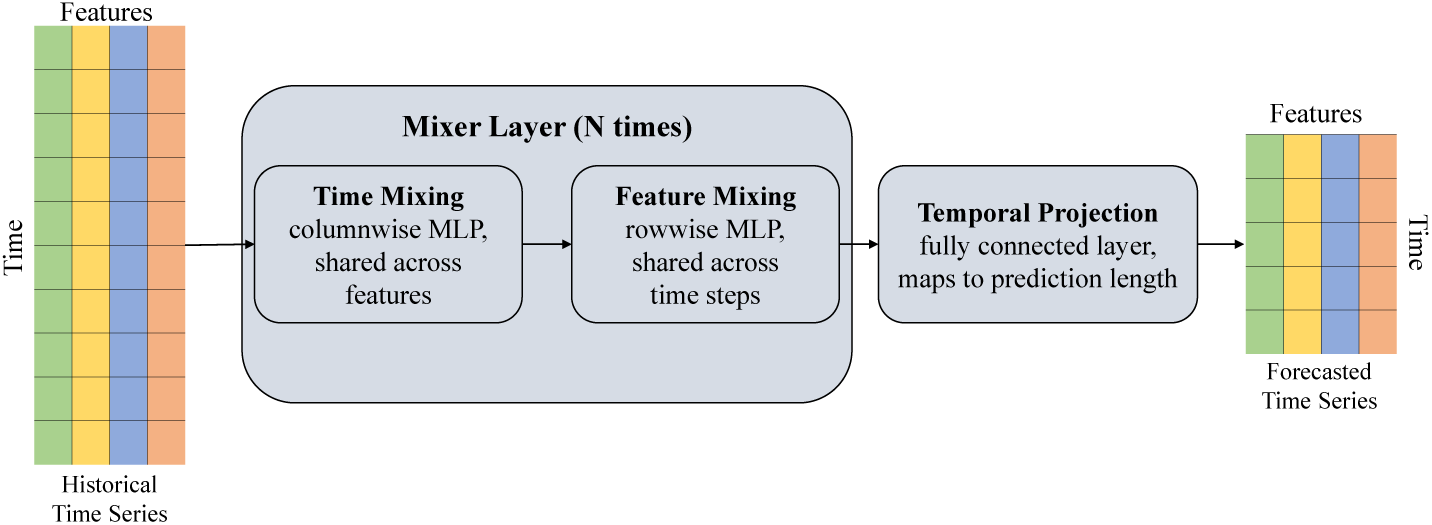
Illustration of the TSMixer architecture, which is designed by stacking multi-layer perceptrons (MLPs). The mixing layers are applied repeatedly across the time dimension and the feature dimension to model both temporal patterns and interdependencies.

The implementation and training scheme follows that of LightGBM as described in the previous subsection.

### 3.4 The mean ensemble and reference models

For the Ensemble, the predictive quantiles were obtained as the arithmetic mean of the respective quantiles of the individual forecasts by the member models (LightGBM, TSMixer, and hhh4). This direct approach, also referred to as *Vincentization* (Genest, 1992) was chosen as other methods like the linear pool are not applicable when only few predictive quantiles are available. As the present analyses shall serve as a blueprint for a collaborative platform with quantile-based submissions (see Section 6), we work with this constraint and thus opt for the Vincentization approach.

To put the performance of the different models into perspective, we apply two simple reference models.

- Persistence is an adaptation of a last-observation-carried-forward prediction to our setting with reporting delays. The predictive mean for horizons 1 through 4 is obtained as the predictive mean of the nowcast distribution at horizon 0. A predictive distribution is obtained as a negative binomial distribution with this mean value, and a dispersion parameter estimated via maximum likelihood from the 15 most recent pairs of predictive means and observations (all obtained using the respective previous data snapshots).
- Historical is a simplistic model only taking into account past seasonal patterns. A predictive distribution for a given calendar week is obtained by collecting all available historical values for said calendar week and the two neighboring weeks and subsequently fitting a negative binomial distribution.

Note that the reference models are not included in the mean ensemble.

### 3.5 Evaluation metrics

The primary evaluation metric is the *weighted interval score* (WIS, Bracher et al. 2021), which can be expressed as a sum of pinball losses. For quantiles *q*_1_, *q*_2_, … *q_K_* at levels *τ*_1_ *< τ*_2_ *<* · · · *< τ_K_* ∈ (0, 1) and an observed value *y* it is given by

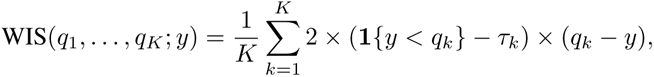

where 1 denotes the indicator function. In our application, we use the previously mentioned levels 2.5%, 10%, 25%, 50%, 75%, 90%, 97.5%. We note that an alternative definition via so-called interval scores exists (hence the name; see Bracher et al. 2021). This display allows for a decomposition into components for forecast dispersion, overprediction, and underprediction, which we will use to enhance the interpretability of performance summary plots.

The WIS is negatively oriented, meaning that lower values are better. It can be seen as a probabilistic extension of the absolute error and approximates the commonly used continuous ranked probability score (CRPS, Gneiting et al. 2005). It is a proper scoring rule, thus incentivizing honest forecasting. To assess forecast calibration separately, the empirical coverage proportions of predictive 50% and 95% prediction intervals are reported.

## 4 Results

### 4.1 Visual inspection of now- and forecasts

We start with a graphical assessment of nowcasts and forecasts. Figure 6 shows now- and forecasts for the total SARI hospitalization incidence (pooled across age groups) issued by the Ensemble at nine different time points. To avoid overplotting, we use two separate panels and display the remaining time points in a set of Supplementary Figures (S2). A detailed illustration of the nowcasts on the aggregate national level can also be found in Figure S3. In Figure 6 at most instances, nowcasts (blue) are closely aligned with the completed data versions (black), but in some cases discrepancies remain (e.g., for the second nowcast in the left panel). The nowcasting also successfully prevents forecasts from following spurious downward trends resulting from reporting delays. Forecasts are mostly well-aligned with the later observed trends, the exception being the first weeks of 2024 (see right panel). Here, the ensemble prediction implies that the peak has already occurred, failing to predict the second and higher peak. Such double peaks in close succession did not occur in any of the previous years, making this aspect hard to predict in a purely data-driven manner. The uncertainty intervals of nowcasts and forecasts are of adequate width to nonetheless cover the observed values in most instances. Especially around the peak, however, they become very wide, meaning that forecasts are less informative in these periods.

**Figure 6:**
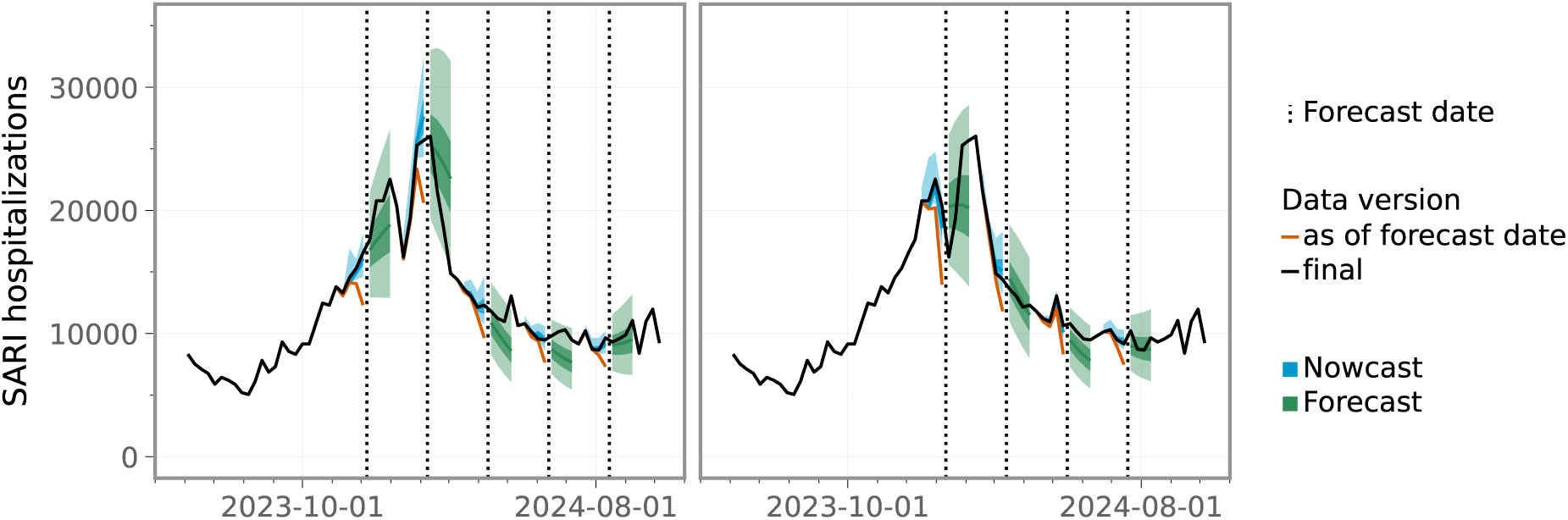
Nowcasts and ensemble forecasts for the total SARI hospitalization incidence (pooled across age groups) at different forecast times. To avoid overplotting, we show the time series twice and overlay it with predictions issued at different times in the two panels. Figures covering all forecast dates are available in Supplementary Figure S2.

Selected predictions from individual models across age groups are displayed in Figure 7. As discussed in Section 2.1, age group 15–34, like some others, displayed unusual patterns in the 2023/24 season. Unlike in previous years, incidence stayed rather high throughout the spring and summer months. The LightGBM model struggles to adjust to this difference and keeps predicting a decline towards the usual levels (a similar pattern arises for TSMixer). The hhh4 model with its simple autoregressive structure is better able to deal with this shift in magnitude. The difficulties of LightGBM and TSMixer are also inherited by the Ensemble. Similar patterns are also found for age group 05–14, and to a lesser degree for ages 35-59, while the remaining age groups have more typical seasonal courses. However, Figure 7 also illustrates some strengths of LightGBM and TSMixer, particularly at the national level (00+) and for older age groups (e.g., 80+). These models accurately capture the sharp decline following the second peak, whereas hhh4 tends to produce more conservative forecasts.

**Figure 7:**
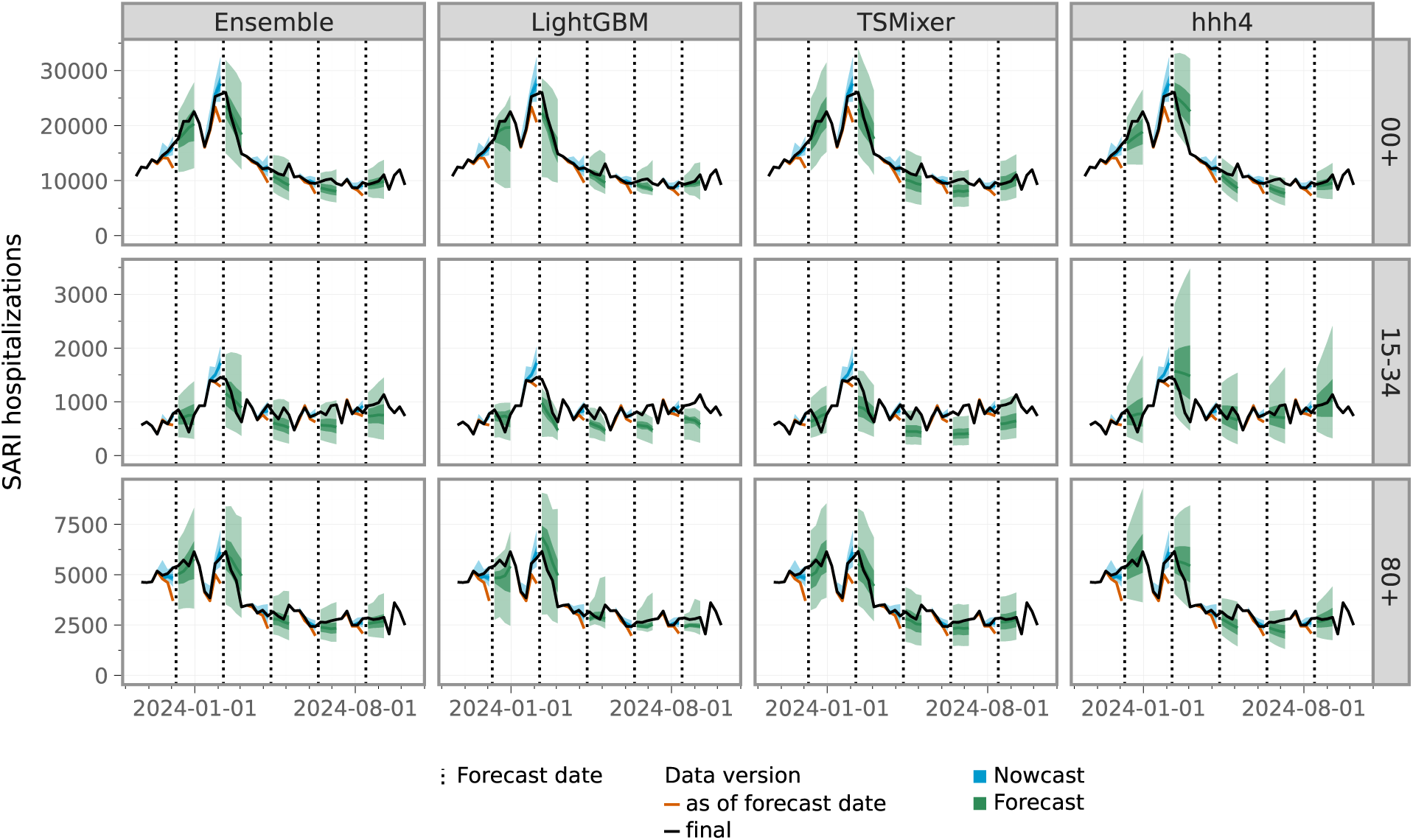
Selected nowcasts and forecasts for the aggregate level 00+ and age groups 15-34 and 80+. To avoid overplotting, predictions for only five forecast times are shown. Figures covering all forecast dates are available for the Ensemble in Supplementary Figure S2.

### 4.2 Formal forecast evaluation

#### 4.2.1 Aggregate-level nowcasts and forecasts

We complement the visual assessment with a more formal evaluation of forecast calibration and score-based performance. Figure 8 summarizes the national-level performance. Average WIS (across forecast dates) as well as the coverage fractions of 50% and 95% prediction intervals are displayed stratified by nowcast/forecast horizon. Little surprisingly, average scores increase with the horizon (i.e., performance decreases). For horizons 1 through 4, all models outperform the Persistence and Historical baseline models (with the exception of TSMixer at horizon 1). The Ensemble outperforms all individual models at all horizons, but the margin to LightGBM and hhh4 is slim for short horizons. Interestingly, for horizon 4 this flips and the TSMixer model achieves performance close to the ensemble. The decomposition of the WIS indicates that LightGBM and TSMixer tend to underpredict, and that this tendency is inherited by the ensemble (this seems to be driven by the fact that the second peak was not anticipated, as well as the untypically high incidences of some age groups late in the season; see previous subsection). The hhh4 and nowcasting models have more balanced components.

**Figure 8:**
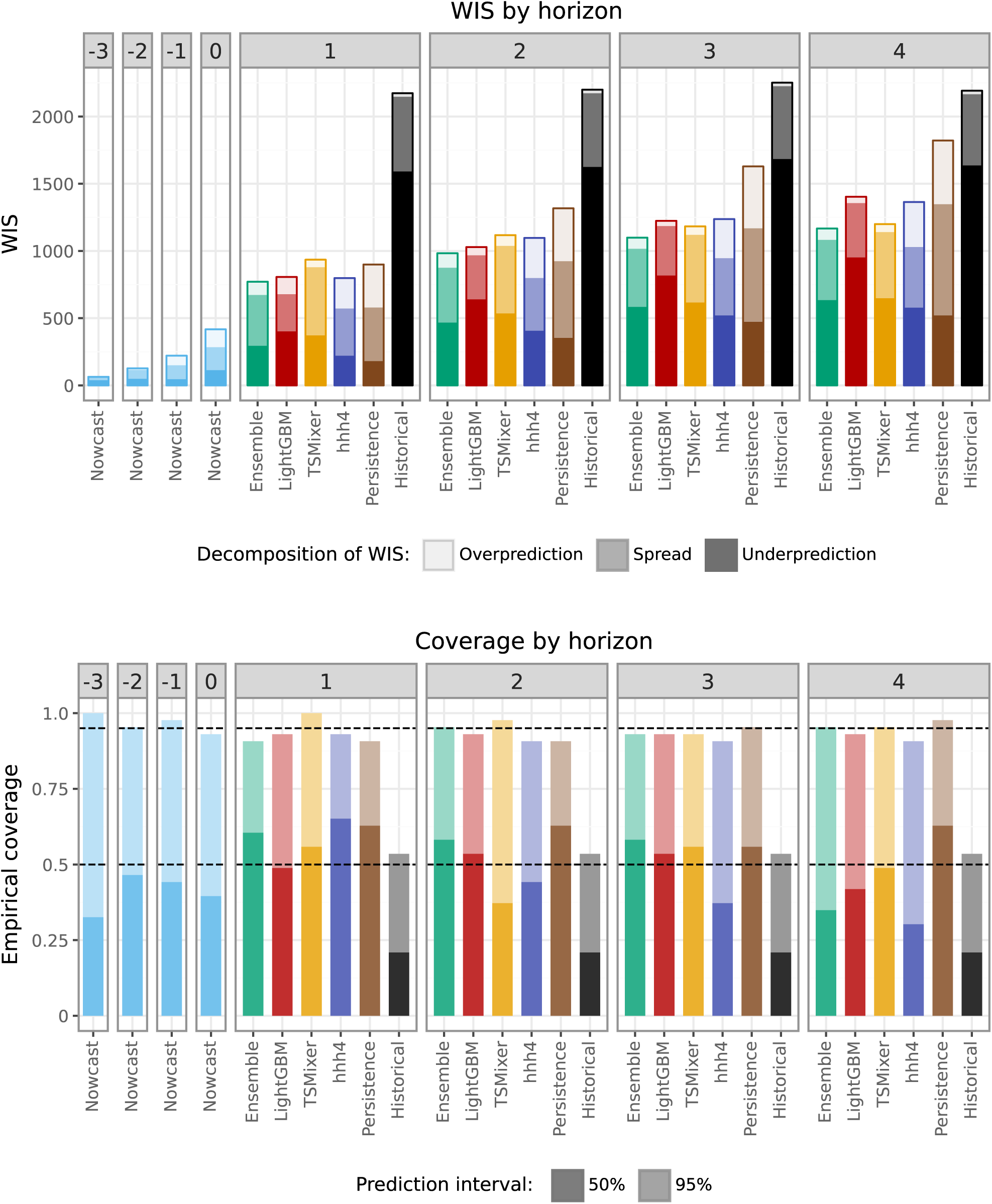
Average WIS values (top) and empirical coverage rates (bottom) achieved by different models for the total SARI hospitalization incidence (pooled across age groups), stratified by nowcast/forecast horizon. The average scores are decomposed into components for overprediction, underprediction, and forecast spread.

A summary plot aggregating results across horizons is available in Supplementary Figure S4 (left panel). While the Ensemble again has a little edge, the three member models LightGBM, TSMixer and hhh4 are roughly on par.

Concerning the interval coverage rates (bottom panel in Figure 8), all models apart from the Historical baseline achieve close-to-nominal coverage.

#### 4.2.2 Age-stratified nowcasts and forecasts

Figure 9 summarizes average results for age-stratified nowcasts and forecasts. The results in terms of average WIS are broadly consistent with those discussed in the previous section, with the ensemble again performing best across horizons and the individual models outperforming the baseline models in almost all instances. The LightGBM and TSMixer models again tend to underpredict, while the hhh4 model features the most dispersed predictions.

**Figure 9:**
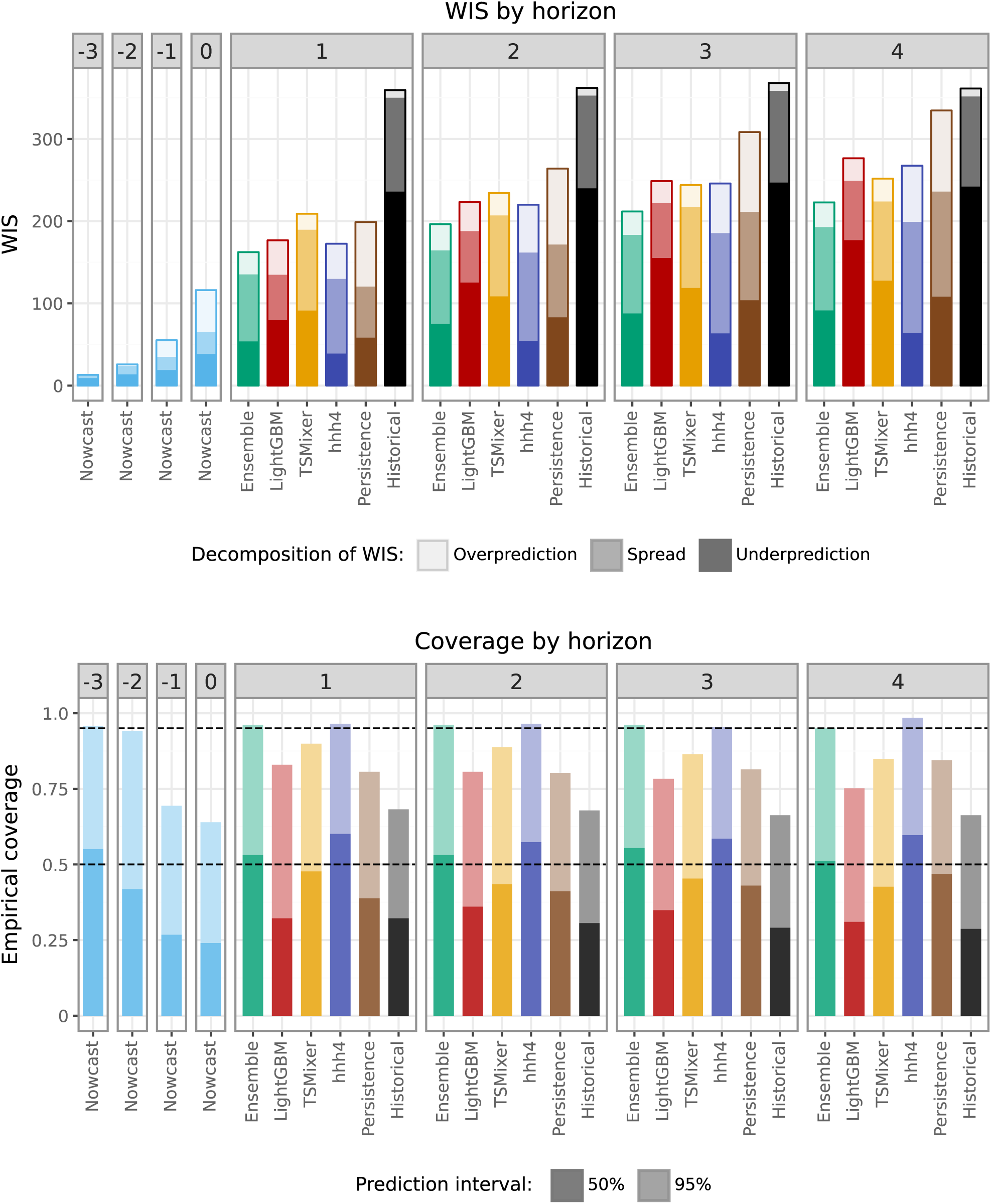
Average WIS values (top) and empirical coverage rates (bottom) achieved by different models for the age-stratified SARI hospitalization incidence. Results are averaged over forecast dates and age groups, and stratified by nowcast/forecast horizon. The average scores are decomposed into components for overprediction, underprediction, and forecast spread.

The WIS stratified by age group (and aggregated by horizon), depicted in Figure 10, reveals that the aforementioned downward bias in LightGBM and TSMixer primarily originates from the age groups 05–14, 15–34, and 35–59. This can be attributed to the unusually high SARI incidence during the evaluation period (Figure 2), which did not follow the typical seasonal decline, as discussed previously. The score-based evaluation also confirms that the hhh4 model performs particularly well in these age groups (in the 15–34 group even slightly outperforming the Ensemble). By contrast, the machine learning approaches had an edge in forecasting older age groups, potentially because they were able to leverage trends in younger age groups as leading indicators for older ones.

**Figure 10:**
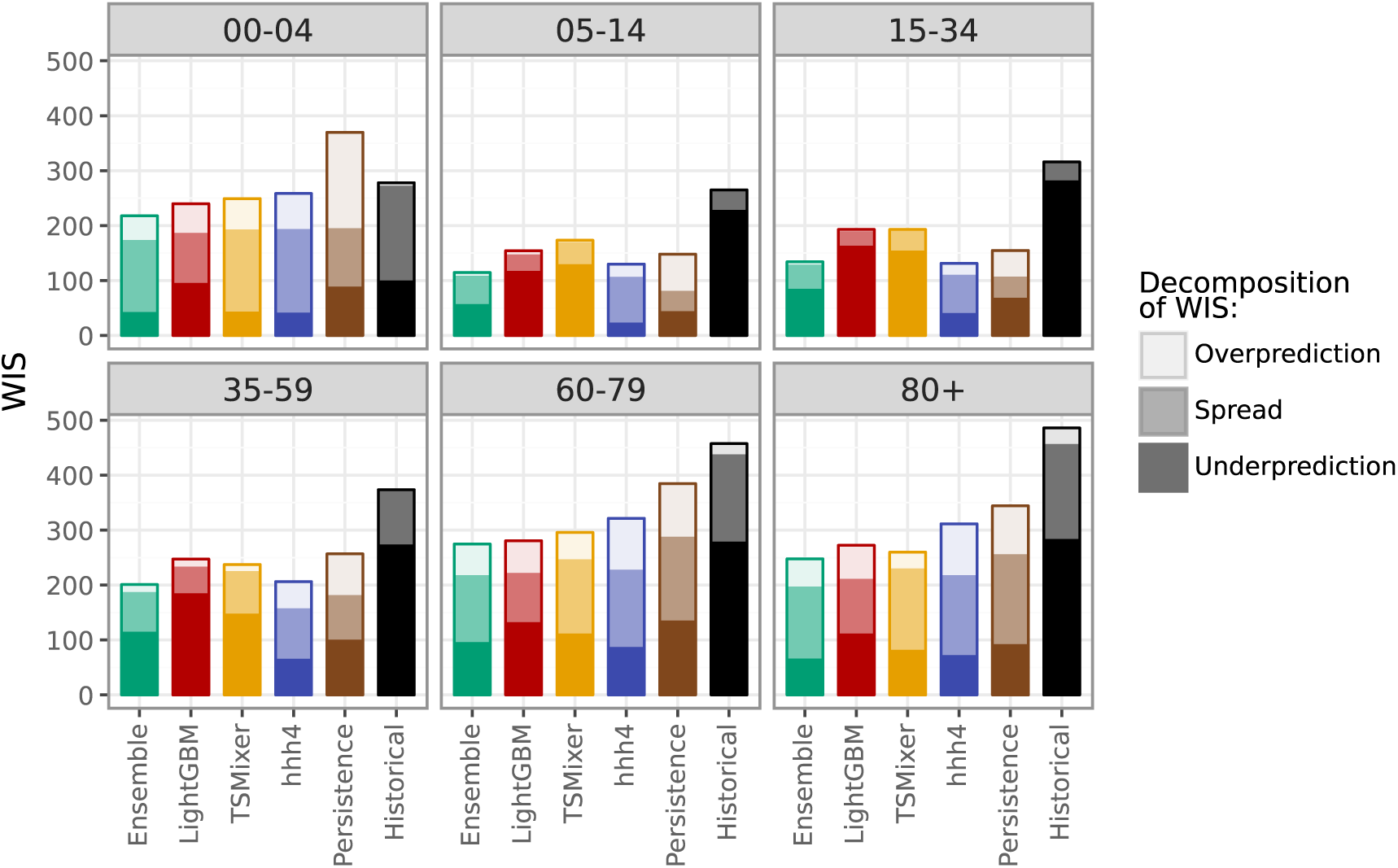
Average WIS by age group, aggregated over forecast dates and horizons. Average scores are decomposed into components for overprediction, underprediction, and forecast spread.

In terms of interval coverage (bottom panel of Figure 9), we observe that the nowcasts for horizons −1 and 0 are considerably overconfident. This is likely a consequence of the fact that only a few historical snapshots of age-stratified data were available, meaning that stratified nowcasts had to be based on aggregate-level snapshots (see Section 3.2). The forecasts from the LightGBM, and to a lesser degree TSMixer models are somewhat overconfident, too. This is not surprising given that the forecasting models take the nowcast as an input. Remarkably, the Ensemble forecast is well-calibrated across horizons and interval levels. This can be explained by the fact that when using Vincentization, the length of the ensemble prediction intervals corresponds to the average length of the member intervals. If the ensemble intervals are centered around a more accurate central tendency (as is often the case), interval coverage rates will tend to increase.

#### 4.2.3 Integration of now- and forecasts

For each of the forecasting methods, we investigate the impact of integrating nowcasts into forecasts and assesss the performance of the chosen implementation route. Thus, instead of including nowcast distributions in the way described in Section 3.2, we apply three alternative strategies.

i. Firstly, we simply ignore the delay problem and use uncorrected incomplete data points to initialize our forecasting models (“Naive”).
ii. Secondly, we discard the last available observation and use only observations that are largely stable, as is common in the literature (Paireau et al., 2022). We still apply the nowcasting procedure to the previous weeks, but this makes little difference in practice (“Discard”).
iii. Lastly, we base forecasts on the final versions of the latest data points, i.e., assess how much forecasts would improve if the reporting system was free of delays. This is a hypothetical setting and not an approach that could be applied in real time (“Oracle”).

Figure 11 summarizes the performance for the total SARI hospitalization incidence under the four considered ways of handling recent data points. Our proposed method of of including the latest data point with a nowcast correction (“Coupling”) yields improvements relative to using uncorrected data (“Naive”) and discarding this data point (“Discard”). This holds especially for short horizons, where the initialization of forecasts is most relevant. In fact, for the hhh4 model, the “Discard” version even works slightly better for horizons 3 and 4. Somewhat surprisingly, when providing forecast models with the final values of recent data points (“Oracle”) rather than nowcasts, performance does not always improve. While for hhh4 it does, for the other models there are minor deteriorations in performance for some horizons. A possible explanation is that initializing the models LightGBM and TSMixer with a nowcast distribution rather than the correct value increases forecast dispersion, which can lead to improved calibration. Corresponding results for age-stratified predictions are shown in Supplementary Figure S6 and are in good agreement with the aggregate-level results.

**Figure 11:**
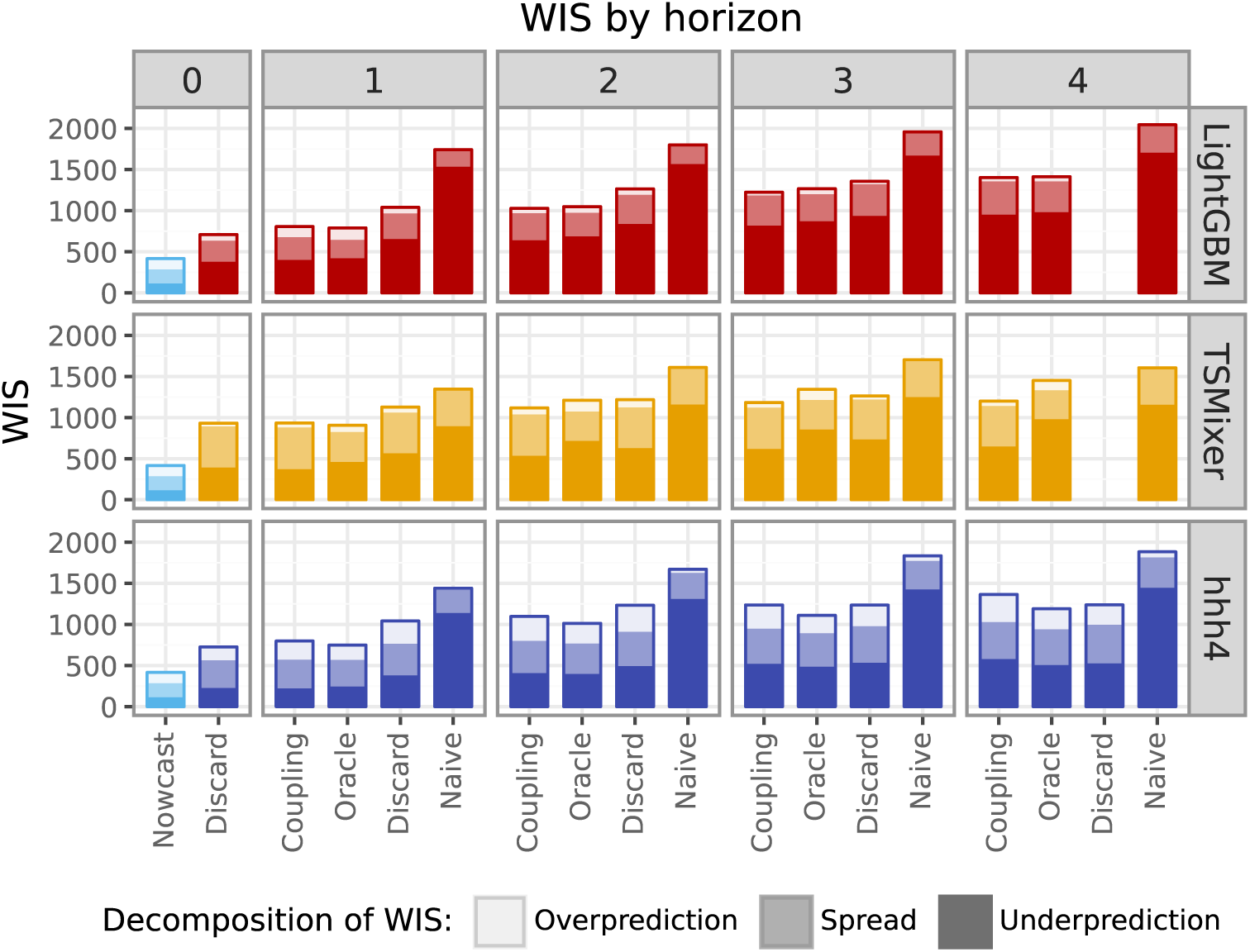
Comparison of forecast performance on the aggregate level resulting from different strategies to handle incomplete recent data. “Coupling” is our main approach described in Section 3.2, i.e. feeding the full nowcast into forecasting models. “Discard” corresponds to discarding the most recent (i.e., most incomplete) data point and treating it like an additional value to be predicted. “Naive” uses the time series as is (with yet incomplete values). “Oracle” is a hypothetical setting where the final versions of the most recent data points are used. It thus enables us to assess the impact of reporting delays on forecast quality.

#### 4.2.4 Robustness checks

Our primary approach consists of including all available data, that comprises the COVID-19 period for training and also uses additional information of ARI to predict SARI incidences. These choices were motivated by hyperparameter tuning, see Supplementary Table S1 where the inclusion of the COVID-19 period and the use of additional ARI data were incorporated into the hyperparameter tuning process.

In this subsection, we study how these implementation decisions impact the predictive performance of each of the forecasting methods. In particular, we exclude the seasons affected by COVID-19 in the training data (see vertical dashed lines in Figure 2) and also omit additional ARI information. In each of the settings, the ML models were re-trained with the corresponding optimal hyperparameters. As before, due to the non-deterministic nature of the training process, we conducted training using 10 different random seeds and averaged the forecasts from these models (i.e., the predictive quantiles at each level) to obtain more robust results.

Supplementary Figure S7 summarizes the effect of excluding data from the COVID-19 period in the training set as well as from omitting ARI incidences as an auxiliary data stream for LightGBM and TSMixer. Using data from the COVID-19 period led to slight improvements in performance for hhh4 and LightGBM. The TSMixer model was very poorly behaved when applied to a reduced data set without the COVID-19 period, indicating that our full time series may already be towards the lower end of the data requirements of this method. The inclusion of the auxiliary time series on outpatient consultations for ARI had only a minor impact on performance, yielding slight improvements for TSMixer and deteriorations for LightGBM.

## 5 Outlook: Prospective evaluation in the RESPINOW Hub

The presented work serves as a blueprint for a broader operational disease nowcasting and forecasting system called the *RESPINOW Hub*. Funded by the German Ministry of Education and Research (BMBF), it is conceived as an open and collaborative system accepting modeling results from multiple research groups. Both conceptually and concerning its technical implementation, it follows the conventions of the Forecast Hub ecosystem (Reich et al., 2022; Cramer et al., 2022; Wolffram et al., 2023). While we here presented a retrospective forecasting exercise based on historical data snapshots, the *RESPINOW Hub* has been collecting real-time predictions on four prediction targets since fall 2024:

- the SARI hospitalization incidence as discussed in the present paper.
- the outpatient consultation incidence for acute respiratory infections (Goerlitz et al., 2021), which in the present work served as an auxiliary data stream.
- the incidence of laboratory-confirmed cases of seasonal influenza as well as respiratory syncytial virus (RSV) as published via the SURVSTAT@RKI 2.0 system (Robert Koch Institute, 2025).

All models presented in this work are also included in the *RESPINOW Hub*. They are complemented by additional nowcasting models and statistical forecasting approaches.This includes in particular a nowcasting approach developed by Robert Koch Institute which will be documented separately. Moreover, mechanistic models for individual pathogens like seasonal influenza are included. The resulting predictions are shared in a weekly rhythm via a dashboard (http://respinowhub.de/) as well as a public GitHub repository (https://github.com/KITmetricslab/ RESPINOW-Hub). An evaluation study on the 2024/25 season has been preregistered (Bracher and Wolffram, 2024), and will shed light on the operational performance of the different models.

## 6 Discussion

We presented and evaluated a multi-model system for nowcasting and short-term forecasting of hospitalizations from severe acute respiratory infections (SARI) in Germany. We addressed this in a modular fashion, where nowcasts were generated in a separate step and subsequently fed into the forecasting models. For short forecast horizons, this led to clear improvements relative to a simpler approach where the most recent data points were used in an uncorrected fashion or simply discarded. Similarly to previous efforts, we found that combined ensemble predictions performed consistently better than individual forecasting models. Forecasts were generally well-calibrated in terms of interval coverage fractions, but in some models as well as the ensemble we observed noteworthy biases in some age groups. Especially the machine learning models LightGBM and TSMixer in these instances seemed to overfit to historical patterns, while the simpler statistical approach hhh4 fared better. In age groups where the seasonal course was closer to historical patterns, however, this model had weaker relative performance.

The good probabilistic calibration of almost all considered models represents a marked difference from results achieved in recent years for COVID-19 cases or deaths (see e.g., Bracher et al. 2021; Cramer et al. 2022). This is surely not due to a sudden improvement in forecasting capacities, but due to the higher predictability of seasonal disease dynamics. Unlike in COVID-19 forecasting, social dynamics and intervention measures were likely no major drivers during the test period. Also, reporting practices were considerably more stable than for most COVID-19 indicators.

Our analyses of forecast performance across horizons and age groups indicate that our three stand-alone models have differing strengths and weaknesses. This *ensemble diversity* is often seen as a key feature for good ensemble performance (DelSole et al., 2014). Especially during the COVID-19 pandemic, collaborative forecasting projects featured considerably more models (the largest effort likely being Cramer et al. 2022 with more than 100 models). This level of effort is unrealistic (and undesirable) outside of times of major crisis. How many models need to be run in order to achieve robust ensemble performance is subject to current research. Fox et al. (2024) recommend using four to seven models and find that the gain from additional models diminishes quickly. In the *RESPINOW Hub*, two more independently run models have recently been included for SARI. We hope this will further enhance the robustness of the ensemble, all while keeping the required effort at a sustainable level.

The ongoing *RESPINOW Hub* project will also enable us to address one of the major weaknesses of the present project, which is the risk of hindsight bias. While we made considerable efforts to manage historical data versions correctly and avoid using data that would not have been available in real time, the applied development and evaluation of prediction models is an iterative process. Implicitly some knowledge on characteristics of the test set may thus have diffused into our forecasting approaches. The follow-up evaluation study of real-time forecasts which we preregistered (Bracher and Wolffram, 2024) will enable us to evaluate the developed models without the risk of hindsight bias.

In the present work, we were exclusively concerned with aggregate SARI hospitalizations which are unspecific to the causative agent. For the 2024/25 season, the Robert Koch Institute started releasing stratified data on SARI hospitalizations caused by COVID-19, seasonal influenza, and RSV. These represent highly relevant additional prediction targets and open new avenues for more mechanistic models explicitly reflecting the dynamics of infection and susceptibility. Such a stratified approach may ultimately also lead to improved forecasts of the total SARI hospitalization incidence.

## Data availability

All data used are available from Robert Koch Institute (https://github.com/robert-koch-institut/SARI-Hospitalisierungsinzidenz/) and in processed form at https://github.com/KITmetricslab/RESPINOW-Hub/tree/main/data/icosari/sari.

## Acknowledgements

This research was supported by the German Federal Ministry of Education and Research (BMBF) via the project RESPINOW (grant number: HZI MV2021-012, NMI FKZ031L0298B). D. Wolffram’s contribution was moreover supported by the Helmholtz Association under the joint research school HIDSS4Health – Helmholtz Information and Data Science School for Health. J. Bracher’s contribution was moreover supported by the German Research Foundation (DFG), project 512483310.

We would like to thank Sam Abbott, Davide Hailer and Jan van de Kassteele for helpful discussions.

The RESPINOW Study Group are: Alex Dulovic, Alex Kuhlmann, André Karch, Berit Lange, Carolina Klett-Tammen, Chao Xu, Claudia Denkinger, Cornelia Gottschick, Daniel Wolffram, Felix Guenther, Florian Marx, Isti Rodiah, Johannes Bracher, Laura-Inés Boehler, Manuela Harries, Melanie Schienle, Michael Böhm, Nicole Schneiderhan-Marra, Nils Bardeck, Olga Hovardovska, Patrick Marsall, Philipp Dönges, Rafael Mikolajczyk, Rolf Kaiser, Sebastian Contreras, Torben Heinsohn, Tyll Krüger, Ulrich Reinacher, Veronika K. Jaeger, Viola Priesemann, Wolfgang Bock.

## A Details on methods

### A.1 Details on hyperparameter tuning

**Table S1:** Hyperparameter Spaces for Tuning.

| LightGBM |  | TSMixer |  |
| --- | --- | --- | --- |
| Parameter | Values | Parameter | Values |
| colsample_bytree | {0.8} | activation | {ReLU} |
| learning_rate | {0.01, 0.05, 0.1} | batch_size | {32} |
| max_bin | {1024, 2048} | dropout | {0.2} |
| max_depth | {-1} | ff_size | {32, 64} |
| min_child_samples | {10, 20, 40} | hidden_size | {32, 64} |
| min_split_gain | {0} | n_epochs | {500, 1000} |
| n_estimators | {500, 1000} | norm_type | {TimeBatchNorm2d} |
| num_leaves | {20, 31, 40} | normalize_before | {false} |
| reg_alpha | {0, 0.5, 1} | num_blocks | {4, 6} |
| reg_lambda | {0, 0.5, 1} | optimizer | {AdamW} |
| sample_weight | {linear, no-covid} | lr | {0.0005, 0.001, 0.005} |
| subsample | {0.8} | weight_decay | {0, 0.001, 0.0001} |
| subsample_freq | {1} | sample_weight | {linear, no-covid} |
| use_covariates | {true, false} | use_covariates | {true, false} |
| Number of combinations: | 3889 | Number of combinations: | 576 |
| Runtime: | 3 days | Runtime: | 6 days |

**Table S2:** Optimal LightGBM hyperparameters for different settings, determined by tuning on the validation period.

LightGBM
| Parameter | All Data | No Covariates | No Covid |
| --- | --- | --- | --- |
| colsample_bytree | 0.8 | 0.8 | 0.8 |
| learning_rate | 0.1 | 0.01 | 0.1 |
| lags | 8 | 8 | 8 |
| lags_future_covariates | [0, 1] | [0, 1] | [0, 1] |
| max_bin | 1024 | 2048 | 2048 |
| max_depth | -1 | -1 | -1 |
| min_child_samples | 10 | 40 | 10 |
| min_split_gain | 0 | 0 | 0 |
| n_estimators | 500 | 1000 | 500 |
| num_leaves | 40 | 40 | 31 |
| reg_alpha | 0.5 | 0 | 0.5 |
| reg_lambda | 0.5 | 0.5 | 0.5 |
| sample_weight | linear | linear | no-covid |
| subsample | 0.8 | 0.8 | 0.8 |
| subsample_freq | 1 | 1 | 1 |
| use_covariates | true | false | true |
| WIS (validation) | 447.06 | 453.60 | 448.50 |

**Table S3:** Optimal TSMixer hyperparameters for different settings, determined by tuning on the validation period.

| Parameter | All data | With Covariates | No Covid |
| --- | --- | --- | --- |
| activation | ReLU | ReLU | ReLU |
| batch_size | 32 | 32 | 32 |
| dropout | 0.2 | 0.2 | 0.2 |
| ff_size | 64 | 64 | 32 |
| hidden_size | 64 | 32 | 32 |
| input_chunk_length | 8 | 8 | 8 |
| normalize_before | false | false | false |
| norm_type | TimeBatchNorm2d | TimeBatchNorm2d | TimeBatchNorm2d |
| num_blocks | 4 | 4 | 4 |
| n_epochs | 1000 | 500 | 500 |
| optimizer | AdamW | AdamW | AdamW |
| lr | 0.0005 | 0.001 | 0.005 |
| weight_decay | 0.0001 | 0.001 | 0.0001 |
| subsample_freq | 1 | 1 | 1 |
| use_covariates | false | true | false |
| WIS (validation) | 356.92 | 359.20 | 432.69 |

## B The ARI data set

The *Arbeitsgemeinschaft Influenza* sentinel surveillance system (Goerlitz et al., 2021) consists of more than 600 general practitioners, who voluntarily provide information on the number of consultations for respiratory infections. Reporting is done directly to RKI either electronically (SEED-ARE) or by fax. We use the consultation incidence for acute respiratory infections (ARI; ICD-10 codes J00 – J22, B34.9 and J44.0) per 100,000 inhabitants. This indicator is not specific to one pathogen and thus forms part of syndromic surveillance. Data are in principle available per age group and region (with certain pairs of German states merged).

**Figure S1:**
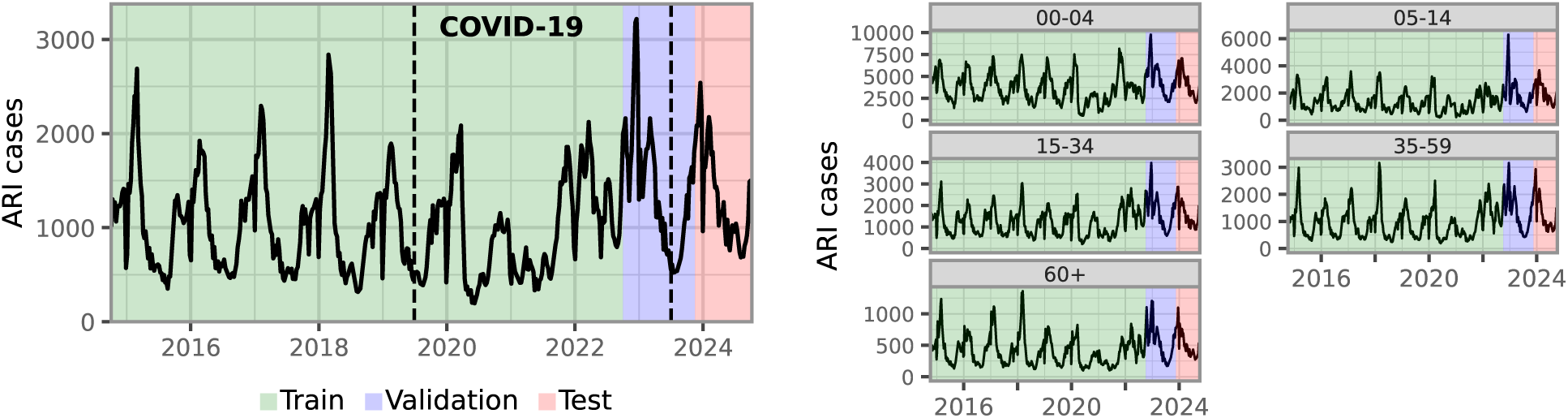
Time series of weekly ARI cases in Germany, 2014–2024. Colors indicate the split of the data into training, validation, and test data. The portion labeled “COVID-19” is only included in the training set for part of our model specifications.

## C Supplementary Figures

**Figure S2:**
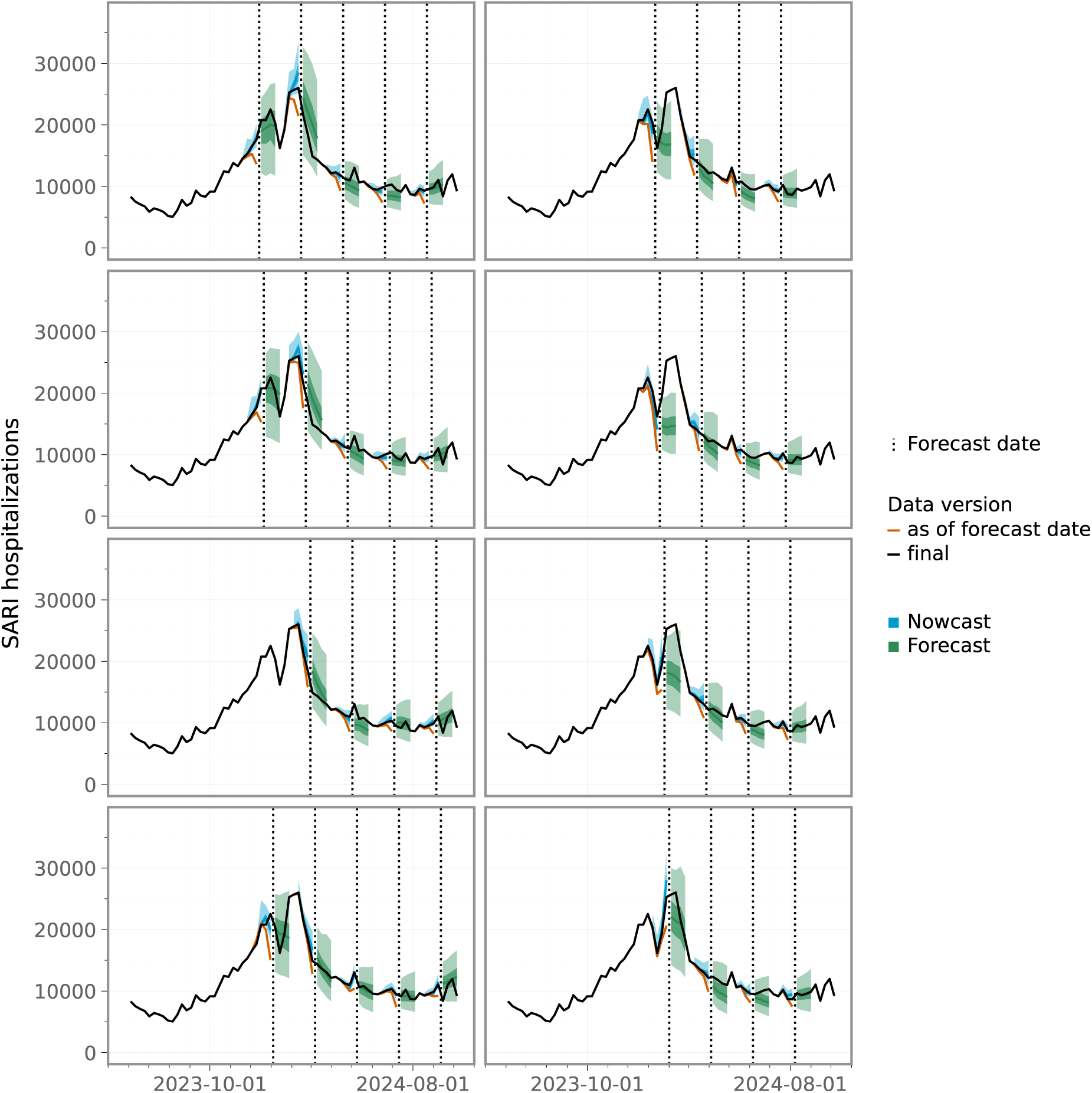
Nowcasts and ensemble forecasts for the total SARI hospitalization incidence (pooled across age groups) at different forecast times. To avoid overplotting, the predictions are displayed in multiple panels.

**Figure S3:**
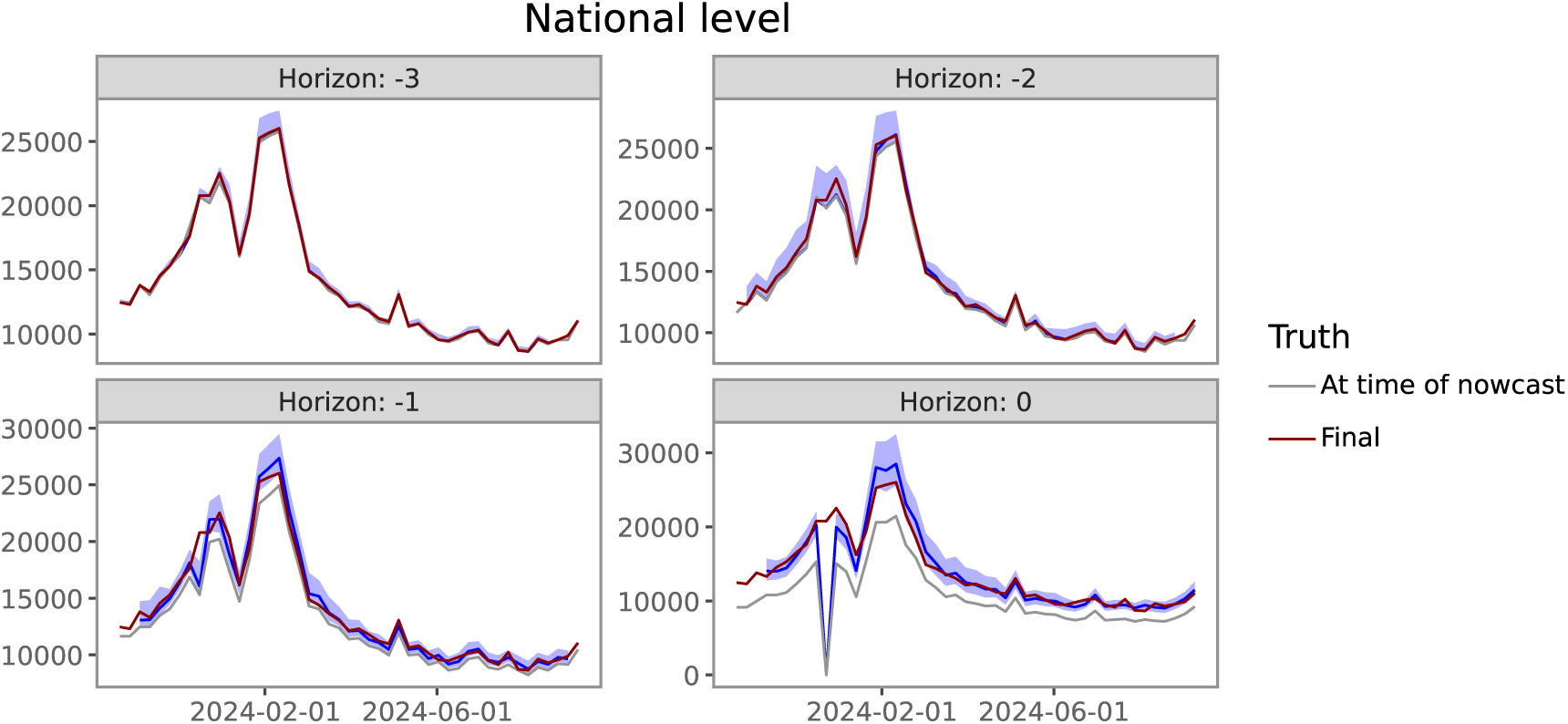
Nowcasts (blue) for the total SARI hospitalization incidence (pooled across age groups), stratified by the horizon. The final values (after 4 weeks) are shown in red, while the gray line represents the incomplete values available at the time of prediction.

**Figure S4:**
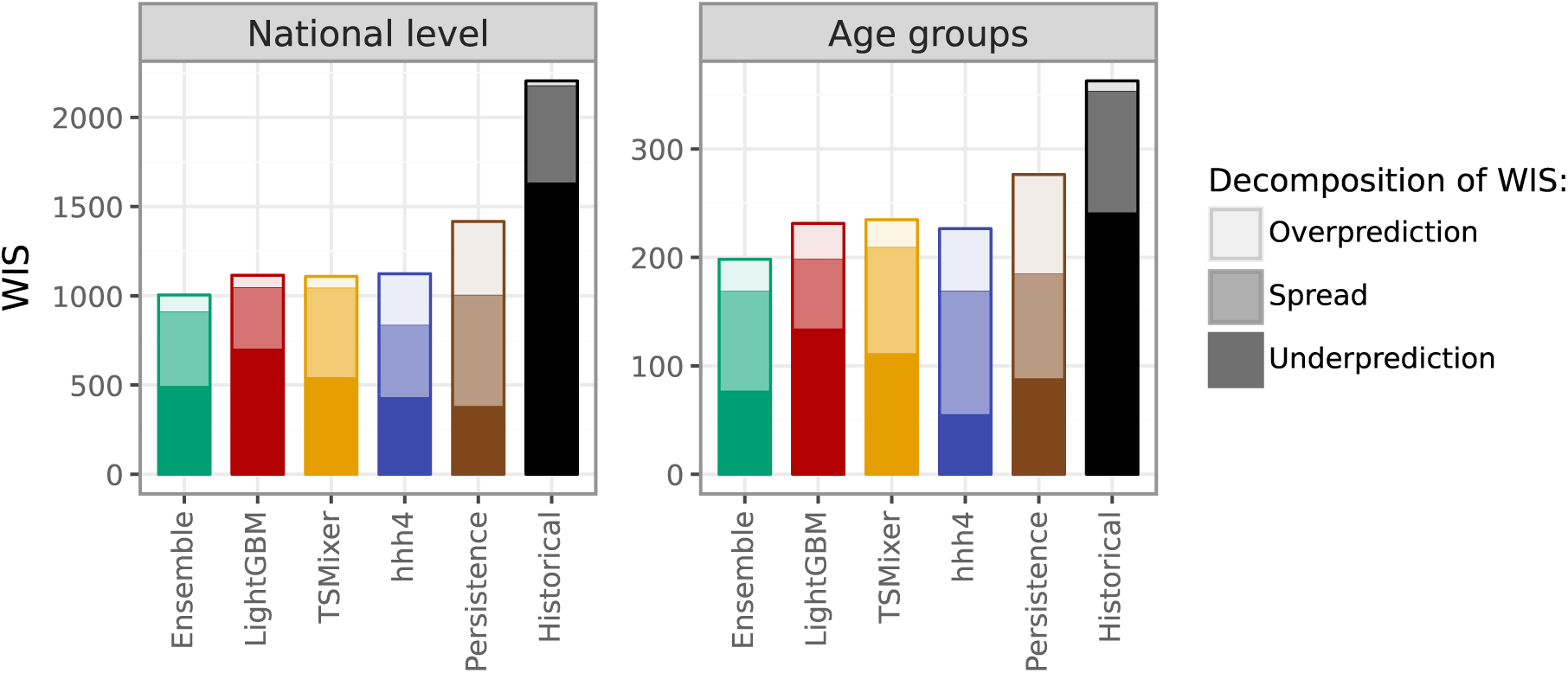
Average WIS on the national level and across age groups. Average scores are decomposed into components for overprediction, underprediction, and forecast spread.

**Figure S5:**
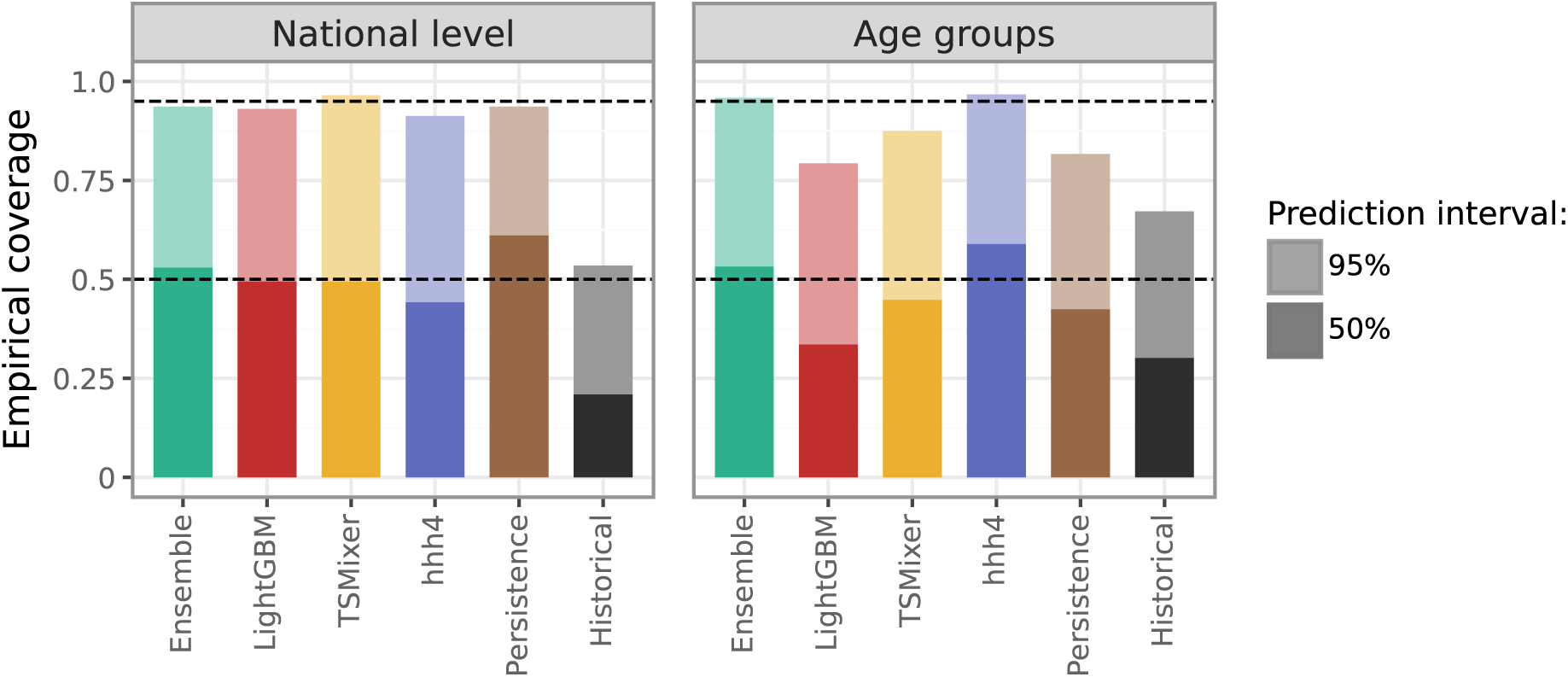
Empirical coverage of the central 50%- and 95%-prediction intervals of different models.

**Figure S6:**
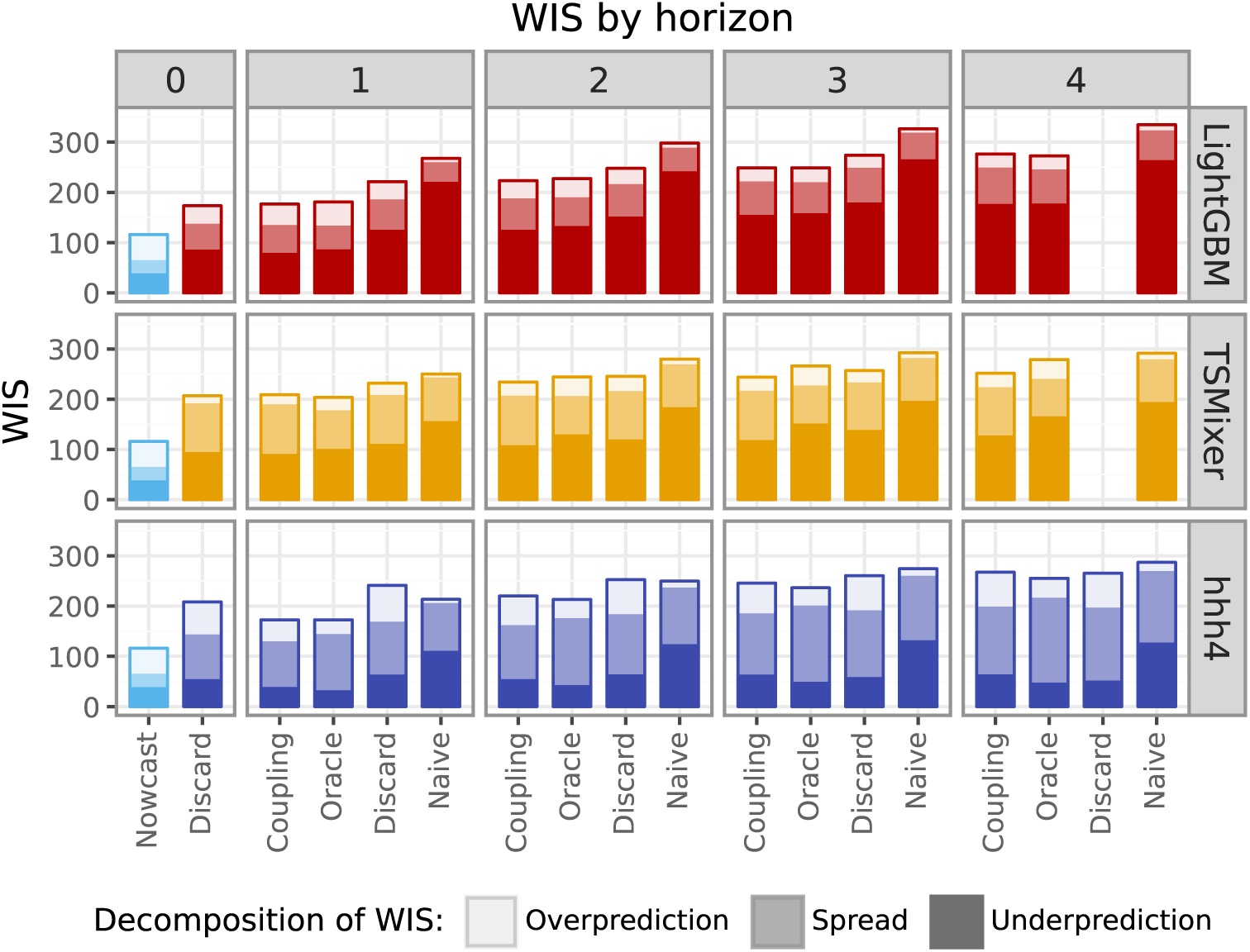
Comparison of forecast performance across age groups resulting from different strategies to handle incomplete recent data. “Coupling” is our main approach described in Section 3.2, i.e. feeding the full nowcast into forecasting models. “Discard” corresponds to discarding the most recent (i.e., most incomplete) data point and treating it like an additional value to be predicted. “Naive” uses the time series as is (with yet incomplete values). “Oracle” is a hypothetical setting where the final versions of the most recent data points are used. It thus enables us to assess the impact of reporting delays on forecast quality.

**Figure S7:**
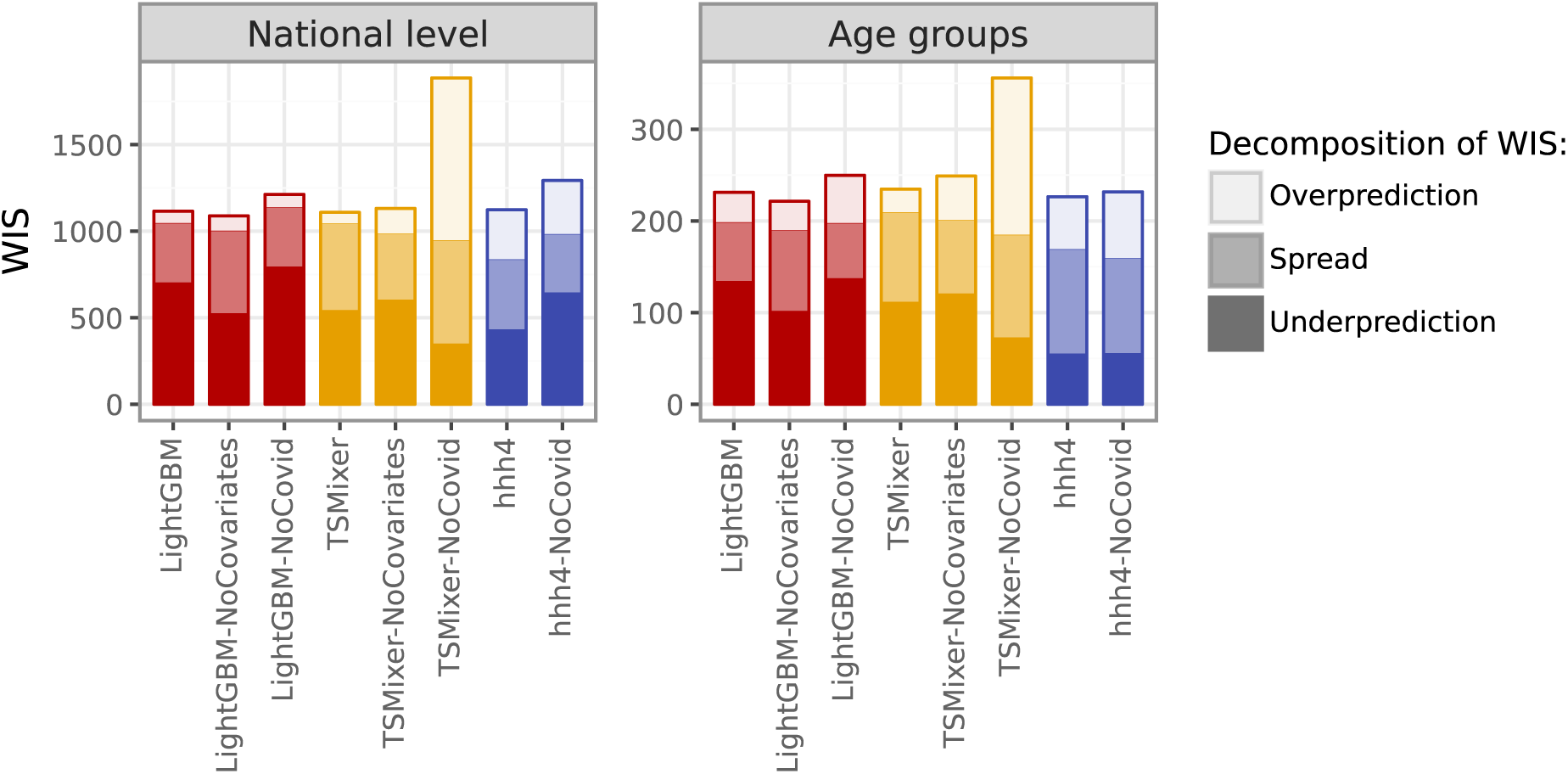
Average WIS (pooled across forecast dates and horizons) for different choices on the training data set. “NoCovid” denotes models where seasons strongly affected by the COVID-19 pandemic (see Figure 2 are excluded). “NoCovariates” denotes models where the auxiliary data set on ARI consultations was removed (note that the hhh4 model does not use this in the first place).

## References

Beesley, L. J., Osthus, D., and Del Valle, S. Y. (2022). Addressing delayed case reporting in infectious disease forecast modeling. PLOS Computational Biology, 18(6):1–26.

Biewald, L. (2020). Experiment tracking with weights and biases. Software available from wandb.com.

Bleichrodt, A., Luo, R., Kirpich, A., and Chowell, G. (2024). Evaluating the forecasting performance of ensemble sub-epidemic frameworks and other time series models for the 2022–2023 mpox epidemic. Royal Society Open Science, 11(7):240248.

Bracher, J. and Held, L. (2022). Endemic-epidemic models with discrete-time serial interval distributions for infectious disease prediction. International Journal of Forecasting, 38(3):1221–1233.

Bracher, J. and Wolffram, D. (2024). Preregistration: Nowcasting and Short-Term Forecasting of Respiratory Infections in Germany, 2024/25. https://osf.io/tgsem/.

Bracher, J., Wolffram, D., Deuschel, J., Görgen, K., Ketterer, J. L., Ullrich, A., Abbott, S., Barbarossa, M. V., Bertsimas, D., Bhatia, S., et al. (2021). A pre-registered short-term forecasting study of COVID-19 in Germany and Poland during the second wave. Nature communications, 12(1):5173.

Buchholz, U., Lehfeld, A.-S., Tolksdorf, K., Cai, W., Reiche, J., Biere, B., Dürrwald, R., and Buda, S. (2023). Respiratory infections in children and adolescents in Germany during the COVID-19 pandemic. Journal of health monitoring, 8(2):20.

Buda, S., Tolksdorf, K., Schuler, E., Kuhlen, R., and Haas, W. (2017). Establishing an ICD-10 code based SARI-surveillance in Germany–description of the system and first results from five recent influenza seasons. BMC public health, 17:1–13.

Chen, S.-A., Li, C.-L., Yoder, N., Arik, S. O., and Pfister, T. (2023). TSMixer: An All-MLP Architecture for Time Series Forecasting. *arXiv preprint arXiv:*2303.06053.

Cramer, E. Y., Ray, E. L., Lopez, V. K., Bracher, J., Brennen, A., Castro Rivadeneira, A. J., Gerding, A., Gneiting, T., House, K. H., Huang, Y., et al. (2022). Evaluation of individual and ensemble probabilistic forecasts of COVID-19 mortality in the United States. Proceedings of the National Academy of Sciences, 119(15):e2113561119.

DelSole, T., Nattala, J., and Tippett, M. K. (2014). Skill improvement from increased ensemble size and model diversity.Geophysical Research Letters, 41(20):7331–7342.

Fox, S. J., Kim, M., Meyers, L. A., Reich, N. G., and Ray, E. L. (2024). Optimizing disease outbreak forecast ensembles. Emerging Infectious Diseases, 30(9):1967.

Genest, C. (1992). Vincentization revisited. The Annals of Statistics, pages 1137–1142.

Gneiting, T., Raftery, A. E., Westveld, A. H., and Goldman, T. (2005). Calibrated probabilistic forecasting using ensemble model output statistics and minimum CRPS estimation. Monthly Weather Review, 133(5):1098–1118.

Goerlitz, L., Tolksdorf, K., Buchholz, U., Prahm, K., Preuß, U., an der Heiden, M., Wolff, T., Dürrwald, R., Nitsche, A., Michel, J., Haas, W., and Buda, S. (2021). Überwachung von COVID-19 durch Erweiterung der etablierten Surveillance für Atemwegsinfektionen. Bundesgesundheitsblatt - Gesundheitsforschung - Gesundheitsschutz, 64(4):1437–1588. 10.1007/s00103-021-03303-2.

Howerton, E., Contamin, L., Mullany, L. C., Qin, M., Reich, N. G., Bents, S., Borchering, R. K., Jung, S.-M., Loo, S. L., Smith, C. P., et al. (2023). Evaluation of the US COVID-19 Scenario Modeling Hub for informing pandemic response under uncertainty. Nature communications, 14(1):7260.

Ke, G., Meng, Q., Finley, T., Wang, T., Chen, W., Ma, W., Ye, Q., and Liu, T.-Y. (2017). LightGBM: A highly efficient gradient boosting decision tree. Advances in neural information processing systems, 30.

Makridakis, S., Spiliotis, E., and Assimakopoulos, V. (2022). M5 accuracy competition: Results, findings, and conclusions. International Journal of Forecasting, 38(4):1346–1364.

Meyer, S., Held, L., and Höhle, M. (2017). Spatio-temporal analysis of epidemic phenomena using the R package surveillance. Journal of Statistical Software, 77(11):1–55.

Paireau, J., Andronico, A., Hozé, N., Layan, M., Crépey, P., Roumagnac, A., Lavielle, M., Boëlle, P.-Y., and Cauchemez, S. (2022). An ensemble model based on early predictors to forecast COVID-19 health care demand in France. Proceedings of the National Academy of Sciences, 119(18):e2103302119.

Reich, N. G., Brooks, L. C., Fox, S. J., Kandula, S., McGowan, C. J., Moore, E., Osthus, D., Ray, E. L., Tushar, A., Yamana, T. K., et al. (2019). A collaborative multiyear, multimodel assessment of seasonal influenza forecasting in the United States. Proceedings of the National Academy of Sciences, 116(8):3146–3154.

Reich, N. G., Lessler, J., Funk, S., Viboud, C., Vespignani, A., Tibshirani, R. J., Shea, K., Schienle, M., Runge, M. C., Rosenfeld, R., et al. (2022). Collaborative hubs: making the most of predictive epidemic modeling. American Journal of Public Health, 112(6):839–842.

Robert Koch Institute (2025). Survstat@rki 2.0. https://survstat.rki.de/Content/Instruction/Content.aspx. accessed 16 January 2025.

Sherratt, K., Gruson, H., Johnson, H., Niehus, R., Prasse, B., Sandmann, F., Deuschel, J., Wolffram, D., Abbott, S., Ullrich, A., et al. (2023). Predictive performance of multi-model ensemble forecasts of COVID-19 across European nations. eLife, 12:e81916.

Tolksdorf, K., Haas, W., Schuler, E., Wieler, L. H., Schilling, J., Hamouda, O., Diercke, M., and Buda, S. (2022). ICD-10 based syndromic surveillance enables robust estimation of burden of severe COVID-19 requiring hospitalization and intensive care treatment. medRxiv.

Wolffram, D., Abbott, S., An der Heiden, M., Funk, S., Günther, F., Hailer, D., Heyder, S., Hotz, T., van de Kassteele, J., Küchenhoff, H., et al. (2023). Collaborative nowcasting of COVID-19 hospitalization incidences in Germany. PLOS Computational Biology, 19(8):e1011394.

